# Accuracy of upper respiratory tract samples to diagnose Mycobacterium tuberculosis: a systematic review and meta-analysis

**DOI:** 10.1101/2022.11.28.22282827

**Authors:** Helen R. Savage, Hannah M Rickman, Rachael M Burke, Maria Lisa Odland, Martina Savio, Beate Ringwald, Luis E Cuevas, Peter MacPherson

**Author notes:** **Corresponding author:** Dr Helen R Savage, Department of Clinical Sciences, Liverpool School of Tropical Medicine, Liverpool, L3 5QA, United Kingdom. Deceased (Luis E Cuevas).

## Abstract

**Structured summary:** 

**Background:** Pulmonary tuberculosis (PTB) due to *Mycobacterium tuberculosis* (Mtb) can be challenging to diagnose because of difficulty obtaining samples, and suboptimal sensitivity of existing tests. We investigated the performance characteristics and diagnostic accuracy of upper respiratory tract tests for diagnosing PTB and hypothesised they would have sufficient accuracy and utility to improve PTB diagnosis.

**Methods:** A systematic review and meta-analysis was conducted by searching MEDLINE, Cinahl, Web of Science, Global Health, and Global Health Archive databases up to 31/01/2021, a second search was conducted for the period 1/1/2021 - 27/5/2022 (subsequently extended to 6/12/2022) to identify studies that reported on the accuracy of upper respiratory tract sampling for TB diagnosis compared to microbiological reference standards. We used a random-effects meta-analysis with a bivariate hierarchical model to estimate pooled sensitivity and specificity, stratified by sampling method. Bias was assessed using QUADAS- 2 criteria. Study registered with PROSPERO (CRD42021262392).

**Findings:** 10,159 titles were screened for inclusion, 274 studies were assessed for full text review, and 71, comprising 119 test comparisons published between 1933 and 2022 were included in the systematic review (53 in meta-analysis). For laryngeal swabs, pooled sensitivity was 57.8% (95% CI 50.5-65.0%), specificity was 93.8% (95% CI 88.4-96.8%) and diagnostic odds ratio (DOR) was 20.7 (95% CI 11.1-38.8). Nasopharyngeal aspirate sensitivity was 65.2% (95% CI 52.0-76.4%), specificity was 97.9% (95% CI 96.0-99.0%) and DOR was 91.0 (95% CI 37.8-218.8). Oral swabs sensitivity was 56.7% (95% CI 44.3-68.2%), specificity was 91.3% (95% CI 81.0-96.3%), and DOR was 13.8 (95% CI 5.6-34.0).

**Interpretation:** Upper respiratory tract sampling holds promise to expand access to TB diagnosis, including for people who can’t produce sputum. Exploring historical methods using modern microbiological techniques may further increase the options for alternative sample types.

Prospective studies are needed to optimise accuracy and utility of sampling methods in clinical practice.

**Funding:** HRS is funded by the MRC through the MRC DTP programme at LSTM [Grant number MR/N013514/1].

**Research in context:** *Evidence before this study:* Globally in 2021, an estimated 4.2 million of 10.6 million people with incident tuberculosis (TB) disease went undiagnosed, emphasising the urgent need for new diagnostic methodologies. Most TB diagnostics are performed on sputum samples, but people who need TB tests are often unable to produce sputum. Upper respiratory tract sampling for TB diagnosis was widely used historically and holds promise to expand non-sputum-based diagnosis.

*Added value of this study:* We systematically reviewed and synthesised through meta-analysis diagnostic accuracy evaluations of upper respiratory tract sampling for TB. Historically, upper respiratory tract sampling for TB diagnosis was commonly used, with 39/71 studies conducted before 1970, although in recent years there has been a resurgence of interest in oral sampling. We show that upper respiratory tract samples have acceptable sensitivity and specificity compared to sputum culture, and, if testing is optimised using newer molecular and culture-based methods, may be capable of meeting WHO target produce profiles.

*Implications of all the available evidence:* Upper respiratory tract sampling methodologies for TB (oral sampling, and sampling from the larynx and nasopharynx) may hold promise to expand access to TB diagnosis, including for people who can’t produce sputum. These sampling strategies can be optimised using modern microbiological techniques to increase access to diagnostics for TB.

## Introduction

Pulmonary disease due to *Mycobacterium tuberculosis (Mtb)* can be challenging to diagnose because of lack of access to testing services, difficulty obtaining samples, and suboptimal sensitivity of tests. In 2021, of the estimated 10.6 million people with incident tuberculosis (TB) over 4.2 million went undiagnosed (1), and of those diagnosed with pulmonary TB (PTB) only 63% were bacteriologically confirmed. Bacterial confirmation is important to ensure correct diagnosis and for identification of drug resistance, so that the most effective treatment regime can be identified (1).

The most common sample used for PTB diagnosis is sputum, which can be tested by smear microscopy, culture, or using nucleic acid amplification tests (NAAT) such as Xpert (Cepheid) or Truenat (Molbio). These methods rely on people producing a sputum sample for analysis. However not everyone being investigated for PTB can produce sputum. For adults in the WHO consolidated guidelines on TB (2) sputum or induced sputum are the recommended sample types for diagnosis of PTB and, if HIV positive, urine using the ALERE-LAM. No alternative non-sputum options are recommended for a microbiological diagnosis of PTB to allow confirmation and resistance testing.

Children commonly do not produce sputum and therefore the WHO recommends alternative sample types for diagnosis of PTB, these include induced sputum, gastric aspirate, gastric lavage, nasopharyngeal aspirate (NPA) and stool (3). Gastric aspirate has a sensitivity of 73% (Xpert MTB/RIF) compared to microbiological reference standard (32% compared to composite reference standard), but is invasive, requires fasting and early morning testing with low caregiver acceptability (2,3). NPA is less invasive however still requires specialist equipment (for suction) and has moderate caregiver acceptability, sensitivity against a microbiological reference standard is 46% (Xpert MTB/RIF, based on four studies) (2,3).

Stool is a newly recommended specimen type in the 2021 update, has high acceptability, and is non-invasive; sensitivity of stool Xpert MTB/RIF is 61% against microbiological reference standard (16% against clinical reference standard) (2,3). However stool testing requires waiting for a bowel movement.

Alternative accessible sample types that can be processed with both existing and novel tests, are patient-centred, and can be collected at the time of consultation, are urgently required.

WHO Target Product Profiles (TPPs) have defined the minimal and optimal diagnostic accuracy standards for tests to diagnose TB in clinical settings, as well as defining desirable characteristics, including using non-sputum samples such as urine, stool, oral mucosal transudates, saliva, exhaled air, or blood from a fingerstick (4). Apart from stool, currently available alternative sample types, such as gastric aspirate or in adults broncho-alveolar lavage are invasive, need fasting and often admission to a hospital with specialist equipment. This can be a barrier to microbiological diagnosis both in terms of availability of the tests, costs, and accessibility.

In Europe in the early 1900s, physicians investigating patients for TB encountered similar concerns and laryngeal swabs were investigated as an alternative sample type for diagnosis of PTB (5–7); however with the decline of TB in Europe this practice fell out of use. More recently, when looking for alternatives to sputum in children, NPA (a procedure where a catheter is inserted into the nostril and suction applied to produce a sample) has been used, and oral mucosa specimens (samples taken by swabbing either the inside of the cheek or the tongue) from South Africa, Moldova and Kenya have successfully been used to diagnose TB (8–10). Sampling of the upper respiratory tract (from the mouth and nose, tonsils, down to the level of the larynx/vocal cords) which is non/minimally invasive, and can be performed quickly as an outpatient, offers a potential alternative to expand access to microbiological TB diagnosis in those that cannot produce sputum.

We therefore set out to systematically appraise the evidence for the performance characteristics and diagnostic accuracy of upper respiratory tract sampling for diagnosing active PTB disease. We hypothesised that upper respiratory tract tests would have sufficient accuracy and utility to increase access to TB diagnosis.

## Methods

We undertook a systematic review and meta-analysis; the published protocol is available online (https://www.crd.york.ac.uk/prospero/display_record.php?RecordID=262392). We developed a search strategy with information specialists at the Liverpool School of Tropical Medicine library (Supplemental Table S1). We searched the following databases up to 31/01/2022, then a further search was carried out for the period 1/1/2021 - 27/5/2022 (subsequently extended to (6/12//2022): MEDLINE; Cinahl; Web of Science; Global Health; and Global Health Archive.

We included studies that evaluated the accuracy of upper respiratory tract sampling (index tests) for a microbiological (culture and nucleic acid amplification tests [NAAT] including automated platforms and laboratory PCR) diagnosis of TB disease, compared to a microbiological reference standard using either sputum or gastric lavage. We included cohort, cross-sectional, and randomised controlled studies (either published in peer-reviewed manuscripts, or as pre-prints) that recruited participants from any community or clinical setting; we placed no restrictions on publication language. We excluded: studies where the index or reference standard used histological or biomarker-based testing; post-mortem studies; studies in non-human animal species; case reports; clinical guidelines; and studies testing for latent TB infection (where active TB disease was not tested for).

Titles and abstracts were imported into a Rayyan.ai database(11), and screened by one reviewer (HRS), with a random subset of 10% checked for agreement by a blinded second reviewer (HMR or RMB). The full text of selected manuscripts was then independently assessed by two reviewers (two of HRS, HMR, RMB) for inclusion; in cases of disagreement, the third reviewer acted as an arbitrator. Risk of bias was assessed using the QUADAS-2 tool (12).

Data were extracted using a piloted extraction form into a study database. Manuscripts published in languages other than English were translated prior to full text review and data extraction.

For each included manuscript, we extracted data on: year; author; country; type of publication; original language; report variable if multiple data sets; study design; setting; population; swab/device used; technique of sampling; site of sampling; number of samples per participant; timing of swabs; swab preparation; transport; storage; method of testing; test cut-off; participants; inclusion criteria; number of samples sets taken; number included in analysis; sex; HIV status; median age; index test positive; index test negative; missing result; reference test positive; reference test negative; missing result; true positive; control population; number of samples sets taken, index test positive, and index test negative.

If a manuscript contained multiple index tests, reference tests or cohorts of patients, each data set was included as a separate report. As participants within a particular study may have been included in more than one index test comparison, we reported the number of sample sets included in the analysis for each test comparison undertaken.

### Data synthesis and analysis

We summarised study characteristics and, separately for each upper respiratory tract sampling methodology, graphed forest plots and calculated the pooled sensitivity and specificity of the index test compared to reference standards using a bivariate hierarchical random effects model, fitted using the lme4 package in R (version 4.2.1) based on the Cochrane handbook for Systematic Reviews of Diagnostic Test Accuracy (13). If studies were of children a second model, using a composite reference standard, including microbiological and clinical cases of PTB was performed (as in the recent WHO consolidated guidelines (2,3)). Definitions of how clinical cases were defined are provided in the Supplementary Material. Data and code to reproduce analysis are available at: https://osf.io/9nuvq/. This study was registered in PROSPERO (CRD42021262392).

### Funding source

HRS is funded by the MRC through the MRC DTP programme at LSTM [Grant number MR/N013514/1]. This research was funded in whole, or in part, by the Wellcome Trust [Grant number 200901/Z/16/Z]. This work was supported by UK Foreign, Commonwealth and Development Office (“Leaving no-one behind: transforming gendered pathways to health for TB”) and partially funded by UK aid from the UK government (to MS, BR, LEC, and PM); however, the views expressed do not necessarily reflect the UK government’s official policies. RMB and HMR are supported by Wellcome PhD Fellowships (203905/Z/16/Z and 225482/Z/22/Z). For the purpose of open access, the author has applied a CC BY public copyright license to any Author Accepted Manuscript version arising from this submission.

The funder of the study had no role in study design, data collection, data analysis, data interpretation, or writing of the report.

## Results

9680 studies were identified during the initial search period up to 31/01/2021, and a further 1364 when extended to 6/12/2022 (Figure 1). We removed 885 duplicates, leaving 10,159 studies that were screened for inclusion. We reviewed the full text of 274 manuscripts.

**Figure 1:**
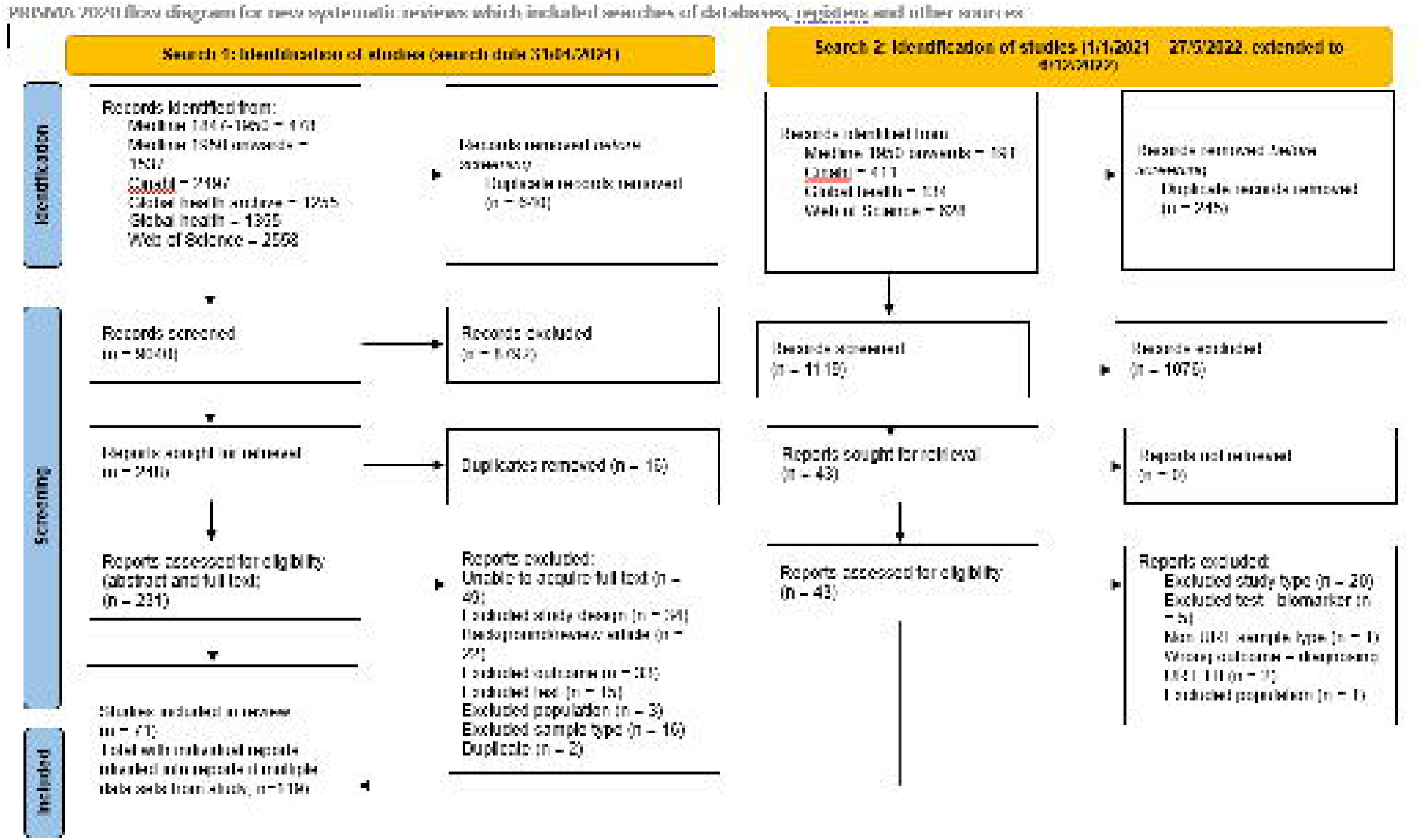
PRISMA 2020 Flow diagram.

Overall, 71 studies were included in the systematic review, comprising 119 reports on index test comparisons; 53 were included in meta-analyses. Full details of excluded studies and reasons for exclusion are in Supplemental Table S2.

From included manuscripts (Table 1; Supplementary Table S3 includes full methodological details and results for each study), we classified types of upper respiratory tract sampling into four groups: laryngeal swabs (32 studies); NPAs (ten studies); oral swabs (18 studies); and other (mouthwash: three studies; nasal swabs: one study; saliva: four studies; other mucosa/dental samples: three studies). Studies were published between 1933 and 2022 from South Africa (eleven studies), Norway (seven studies), UK (seven studies), Peru (four studies), Uganda (seven studies), Canada (three studies), India (three studies), USA (three studies), Australia (two studies), Germany (two studies), Kenya (two studies), and one study from each of Brazil, Chile, China, Czechoslovakia, Denmark, Finland, France, Hungary, Italy, Japan, Malawi, Moldova, Mozambique, Republic of Korea, Spain, South East Asia (not specified), Sweden, Taiwan, Turkey, and Yemen.

**Table 1:**
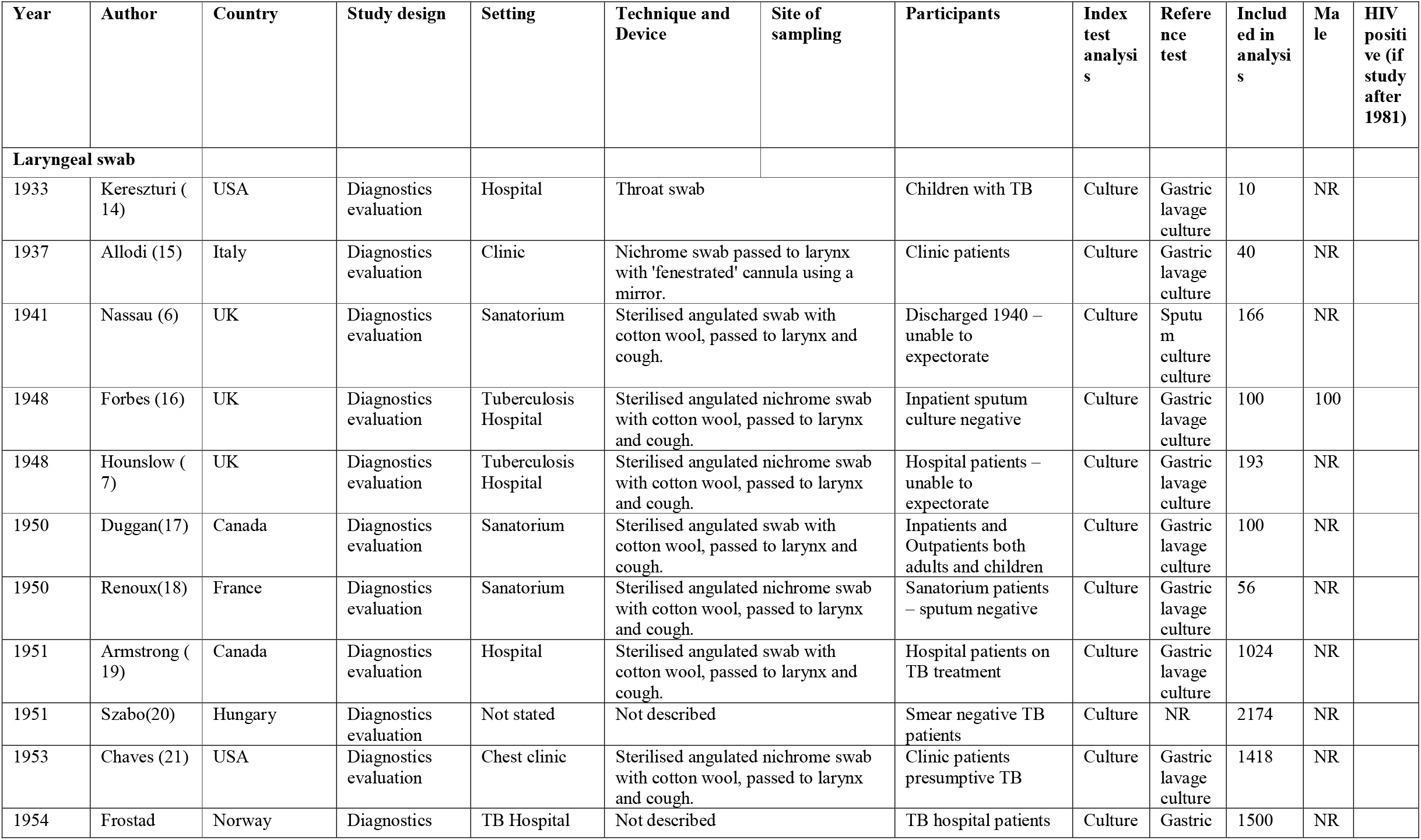
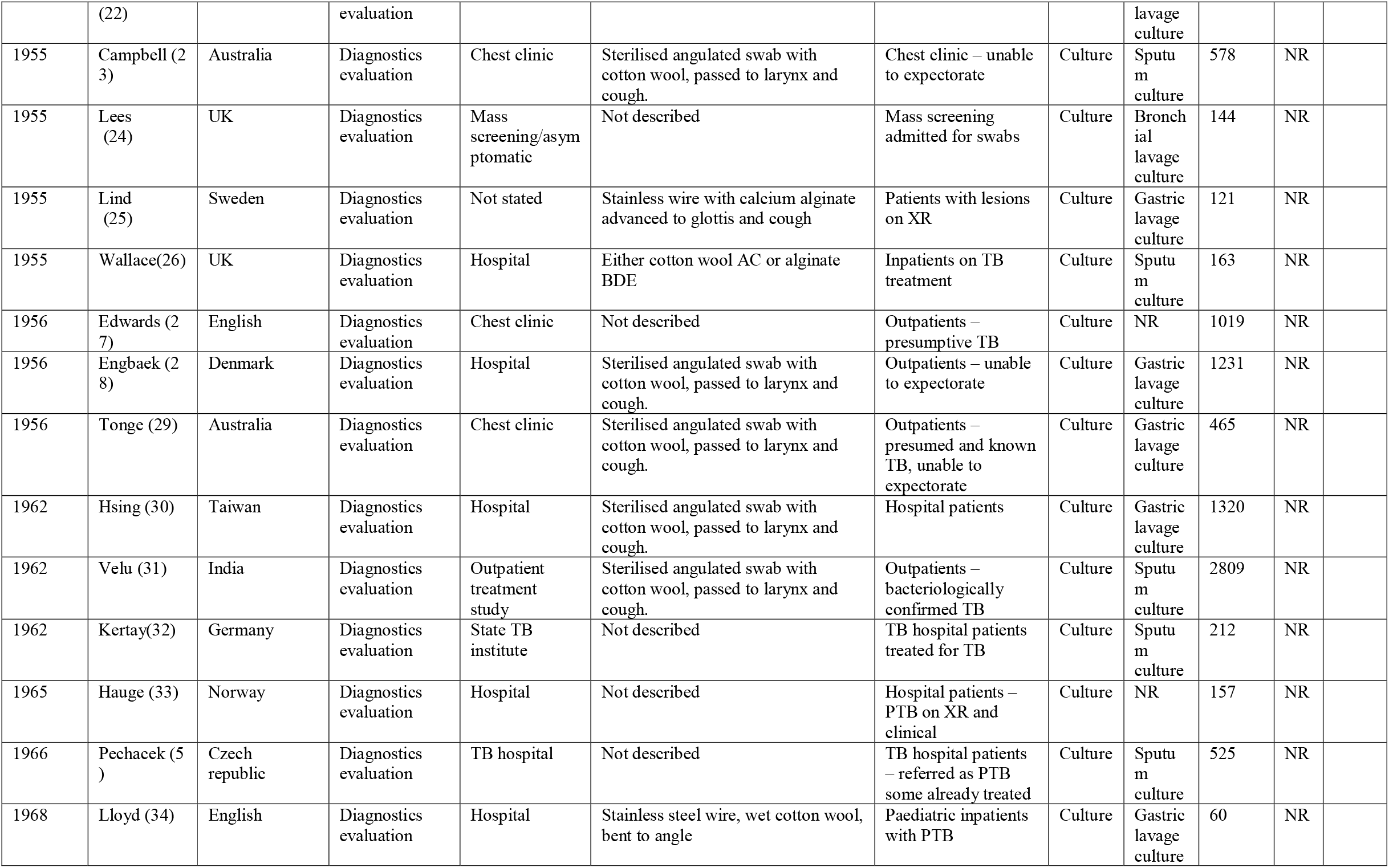
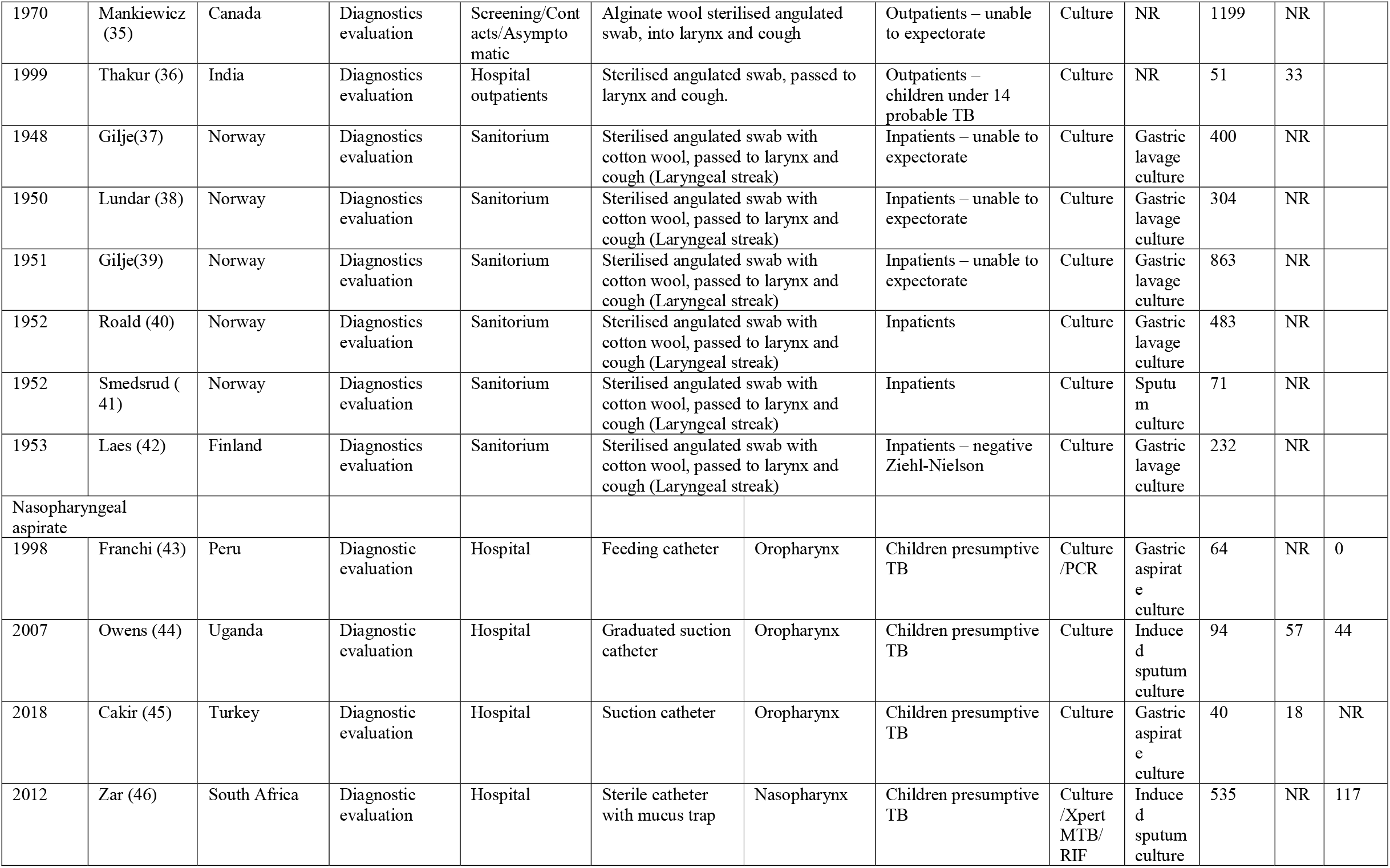
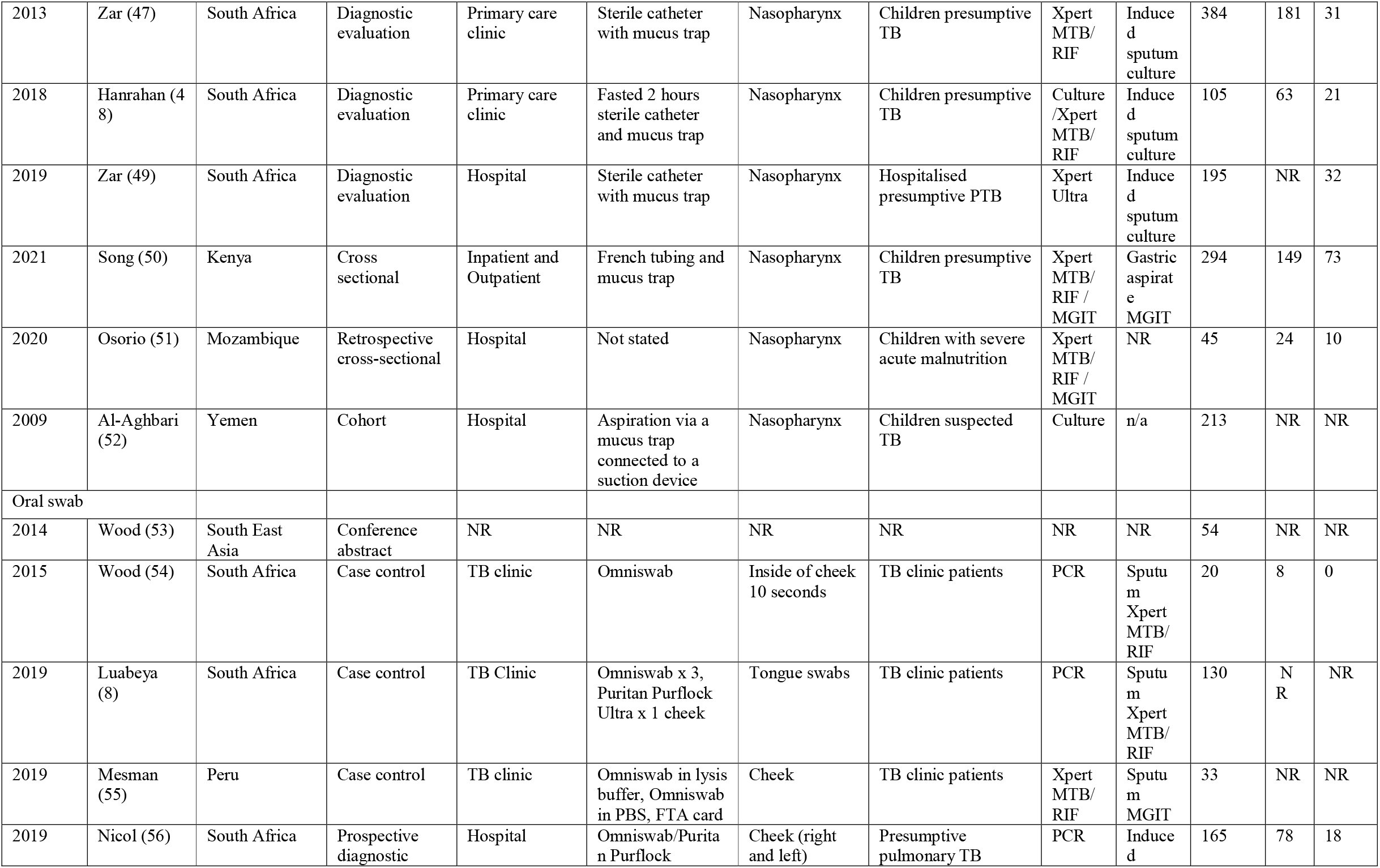
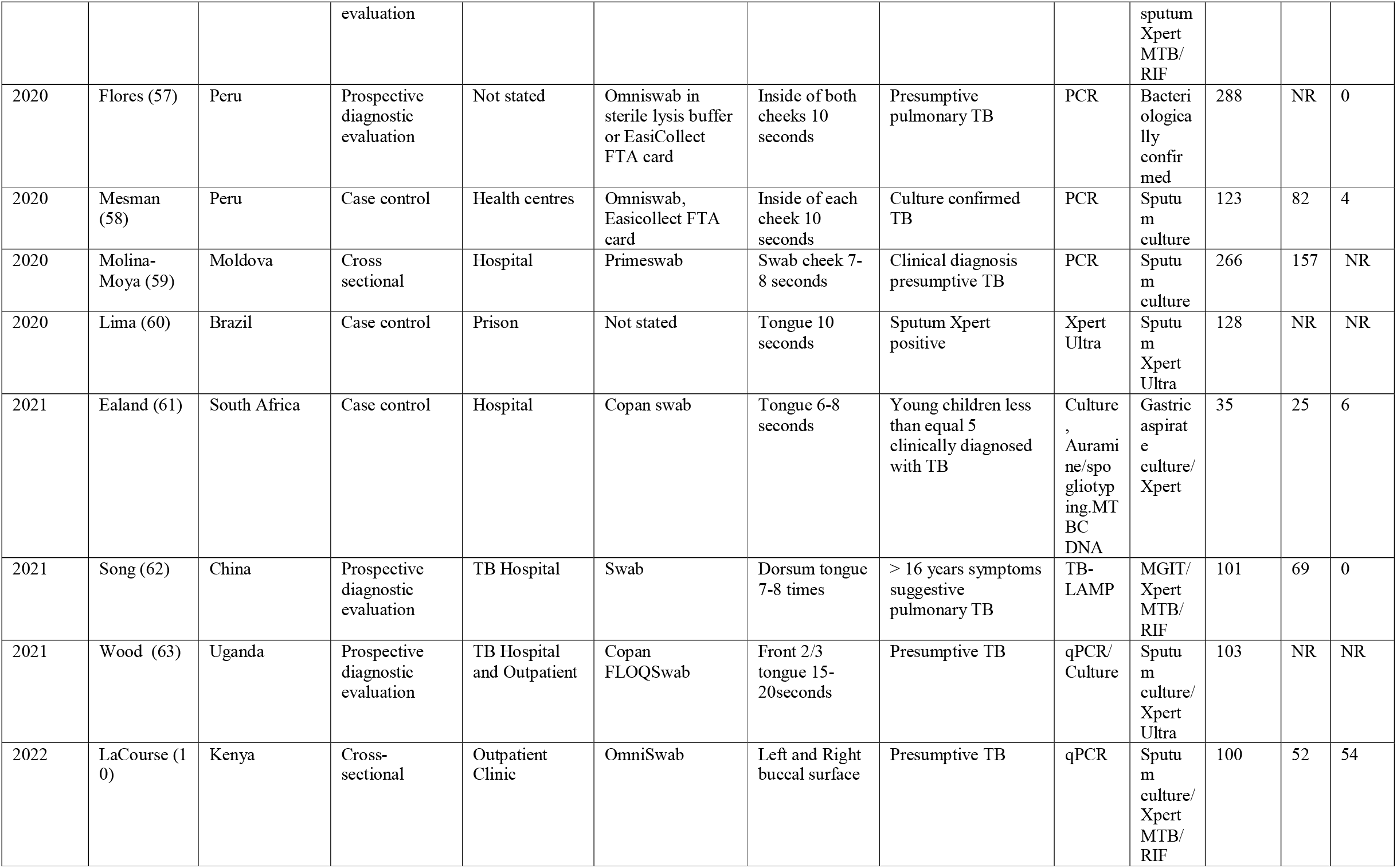
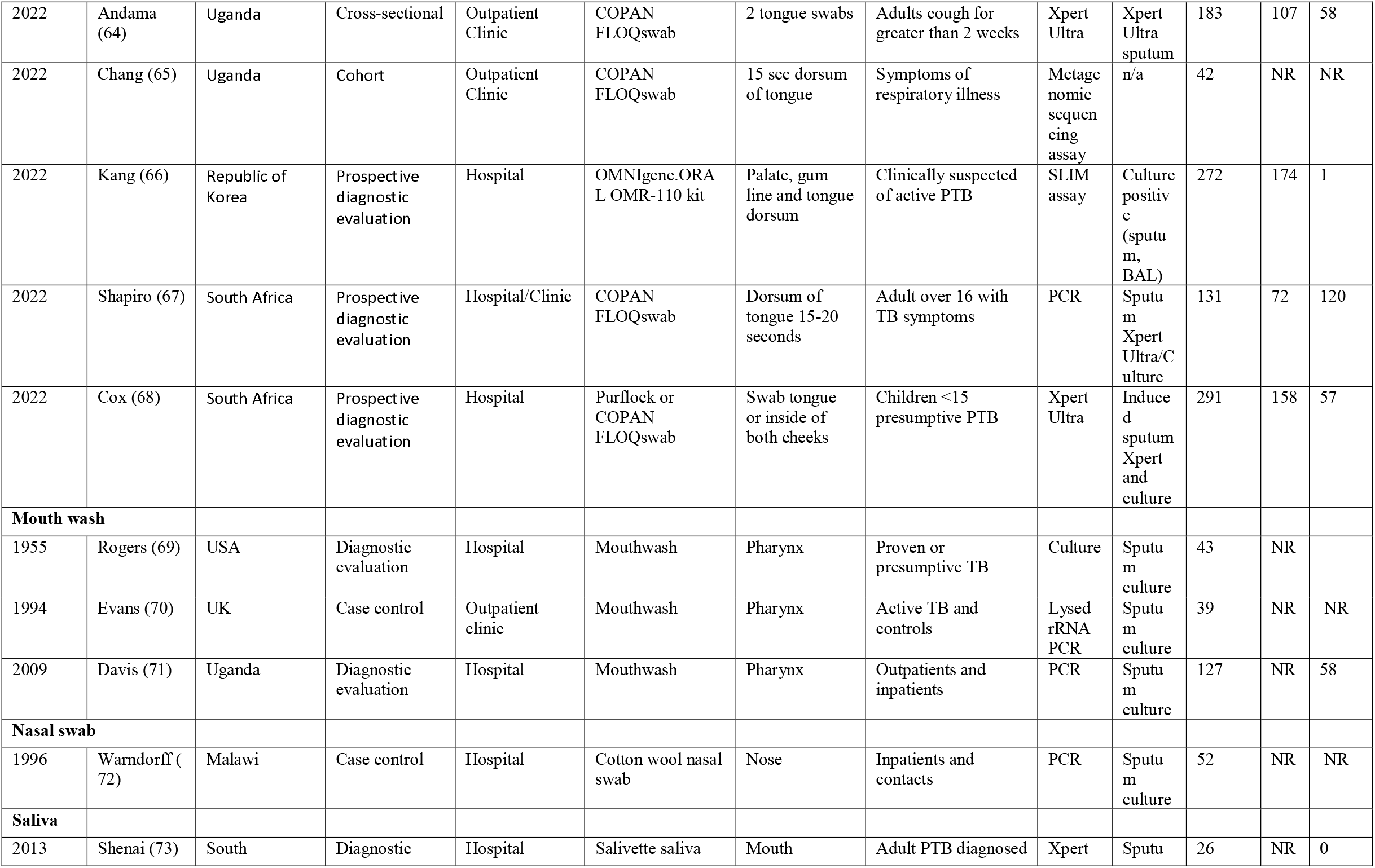
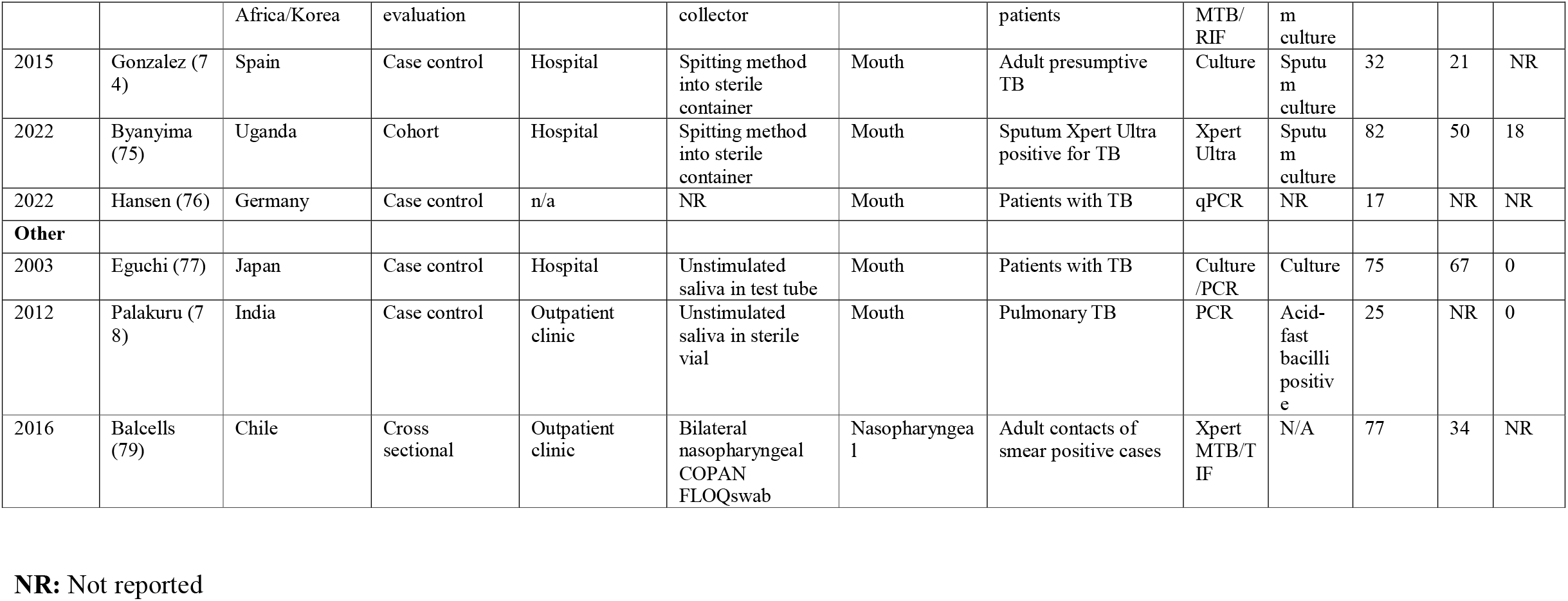
Study Characteristics

Overall, we included data on 24,899 index test samples from participants in hospitals (29 studies); TB sanatoria (nine studies); Chest clinics (nine studies); TB hospitals (eight studies), hospital outpatients (seven studies); primary care clinics (three studies); mass screening/asymptomatic/contact interventions (two studies); outpatient treatment centres (one study), prisons (one study) and studies where the setting was unknown (five studies). Eleven studies were in children aged 16 years or younger. In studies that reported demographic data, 58.2% of participants were male (data from 24 studies) and 19.5% were HIV-positive (with 22 out of 39 studies conducted after 1981 presenting data on HIV prevalence in study participants).

### Laryngeal swabs

Forty-one reports that evaluated the accuracy of laryngeal swabs were included, of which 23 contributed data to the meta-analysis (Supplementary Table S4). Studies were published between 1941 and 1968, with sample sizes ranging from 10 to 2809. A variety of angulated metal swabs with cotton wool or other absorbent materials were used to sample from the larynx, and processed using acidic preparations (Table 1 and Supplemental Table S3).

Samples (both index and reference) were then cultured using methodologies available at the time of the study including modified Petragnani’s medium, Lowenstein-Jensen solid medium, liquid oleic acid albumin media and guinea pig inoculation. In all studies, the reference test used was culture of sputum or gastric lavage fluid (using either expectorated sputum [eight studies] or gastric lavage [15 studies]). Random-effects model estimated sensitivity was 57.8% (95% CI: 50.5-65.0%), specificity 93.8% (95% CI: 88.4-96.8%), diagnostics odds ratio (DOR) 20.7 (95% CI: 11.1-38.8), positive likelihood ratio (PLR) 9.3 (95% CI: 5.1-17.0), and negative likelihood ratio 0.45 (95%CI 0.38-0.53) – Figure 2 (SROC Supplementary Figure S1).

**Figure 2:**
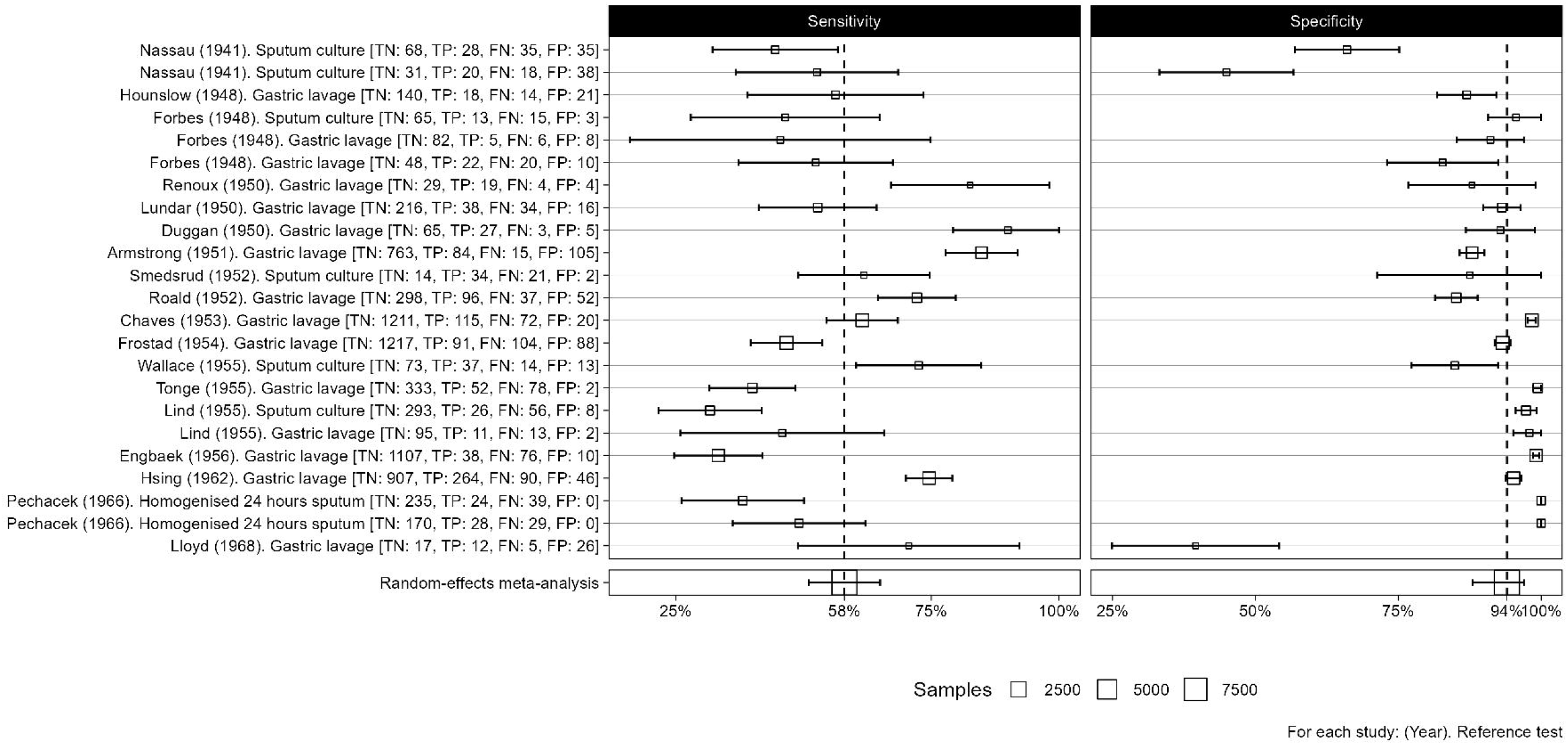
Sensitivity and specificity of laryngeal sampling for active pulmonary tuberculosis, with random effects meta-analysis.

### Nasopharyngeal aspirates

Nine studies that included data on the accuracy of NPA were included, providing 17 test comparisons, of which 10 were included in the meta-analysis. All studies were conducted among children and published between 1998 and 2021, with the number of sample sets ranging from 64 to 535. Multiple index testing methodologies (non-automated culture [four reports], MGIT [one report], Xpert MTB/RIF [three reports], Xpert Ultra [one report], PCR [one report]) were carried out on samples and each was included as a separate comparison. For the microbiological reference test induced sputum culture (6 reports) or gastric aspirate culture (4 reports) were used – Supplementary Table S5. The proportion of children with a microbiologically confirmed diagnosis of PTB ranged from 3% (48) to 38% (43) within the included studies. Model-estimated sensitivity was 65.2% (95%CI 52.0-76.4%), specificity 97.9% (95% CI 96.0-99.0%), DOR 91.0 (95%CI 37.8-218.8), PLR 32.2 (95% CI 15.8-66.1) and NLR 0.35 (95%CI 0.25-0.51) – Figure 3 (SROC, Supplementary Figure 2).

**Figure 3:**
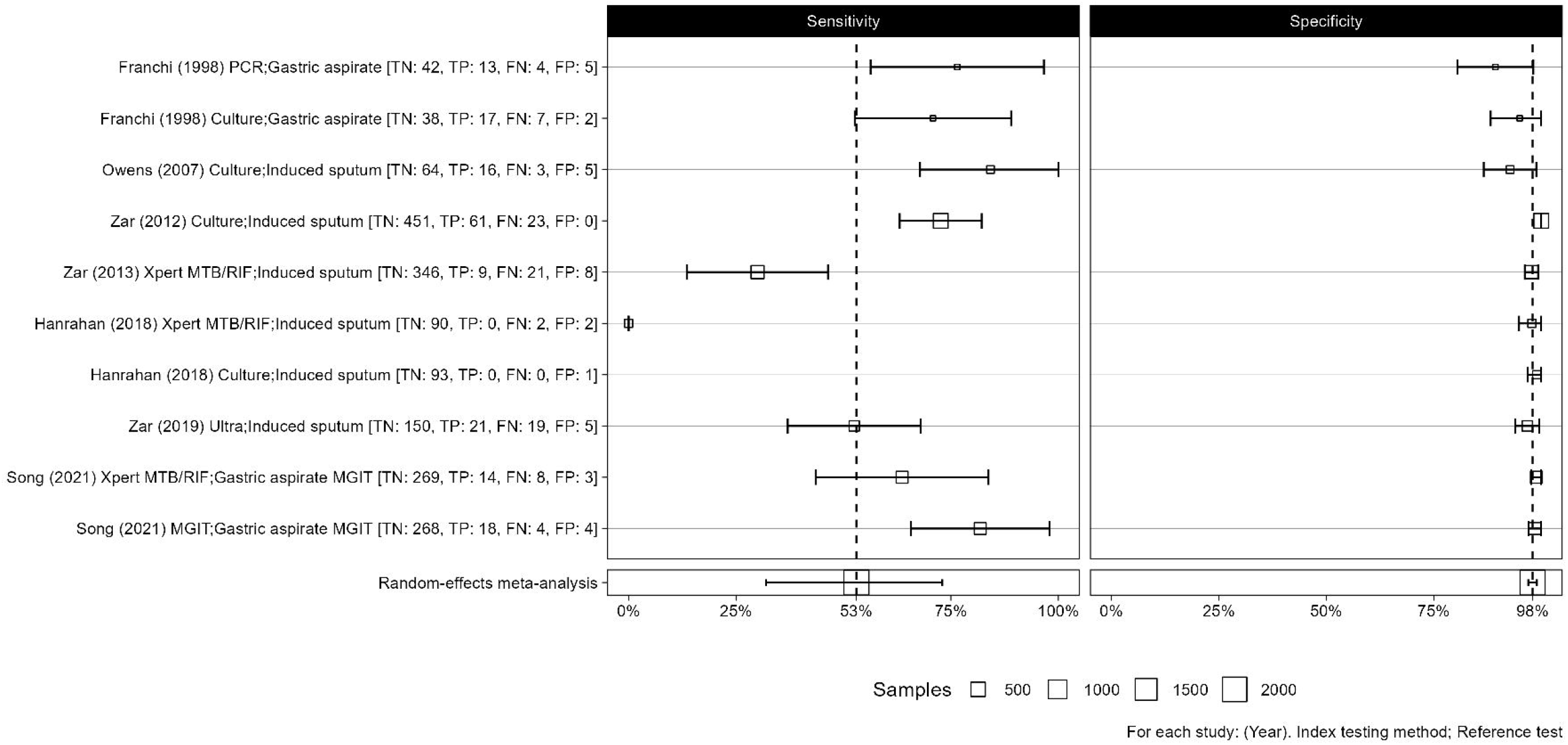
Sensitivity and specificity of nasopharyngeal aspirate for active pulmonary tuberculosis, with microbiological reference standard, with random effects meta-analysis.

A second model was calculated of reports that included study data of a composite reference standard (both microbiological and clinical diagnosis of PTB, see Supplementary Table 6 for inclusion criteria), with 12 comparisons included in the analysis (Supplementary Table S7). The random effects model gave an estimate of sensitivity 9.1% (95%CI 5.6-14.6%), specificity 99.9% (95% CI 93.6-99.9%), DOR 168.4 (95%CI 1.57-17959.1), PLR 153.1 (95% CI 1.43-16343.5) and NLR 0.91 (95%CI 0.87-0.95) – Figure 4 (SROC, Supplementary Figure 3).

**Figure 4:**
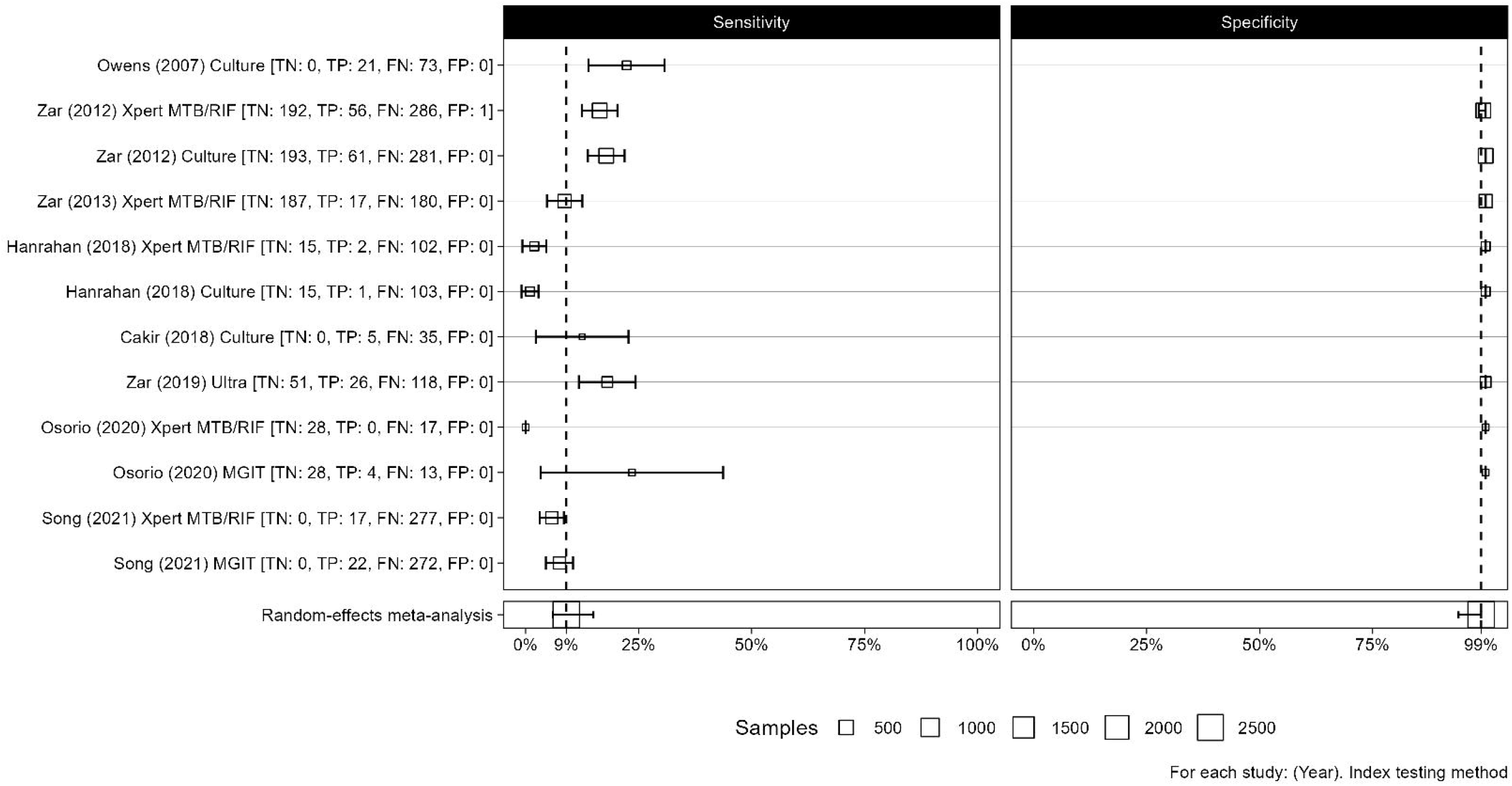
Sensitivity and specificity of nasopharyngeal aspirate for active pulmonary tuberculosis when clinical diagnosis used as a reference standard, with random effects meta-analysis.

### Oral swab

Eighteen studies, with 29 comparisons of oral swab samplings to microbiological reference standards were identified; of these twenty (2015-2022) were included in the meta-analysis.

Ten used PCR for analysis, three used Xpert Ultra, two used culture and one comparison each used Xpert MTB/RIF, TB-LAMP, spoligotyping, auramine smear and SLIM assay. Eight comparisons had sputum culture as reference standard, five used sputum culture and Xpert, two used sputum Xpert alone, four used gastric lavage culture and Xpert and one used sputum or bronchiolar lavage culture (Supplementary Table S8). Seven comparisons included children as participants and thirteen included adults. Pooled sensitivity of oral swab samples was 56.7% (95%CI 44.3-68.2%), specificity 91.3% (95% CI 81.0-96.3%), DOR 13.8 (95%CI 5.6-34.0), PLR 6.54 (95% CI 3.0-14.5) and NLR 0.47 (95%CI 0.36-0.62) – Figure 5.

**Figure 5:**
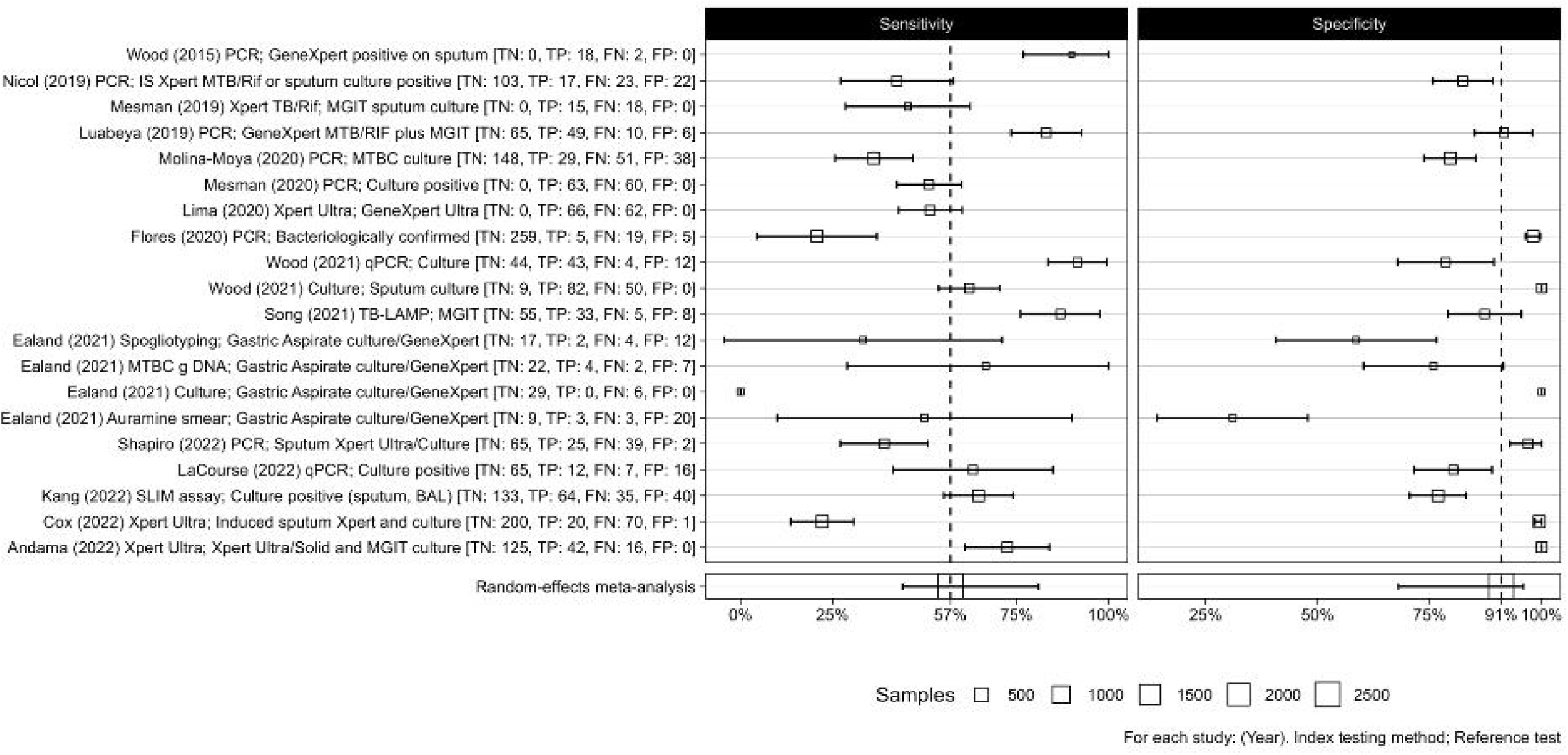
Sensitivity and specificity of oral swab for active pulmonary tuberculosis, with random effects meta-analysis.

### Other sample types

We identified only a small number of studies of alternative sample types, so meta-analysis was not undertaken. Of these three studies compared the accuracy of mouthwash samples, saliva (four studies), oral cavity samples (two studies) and nasopharyngeal swabs (one study) - Supplementary Table S9. A further study on nasal swabs and one on saliva did not contain sufficient information to permit summary of results.

### Risk of bias assessment

The QUADAS-2 tool was used to assess the risk of bias for each study included (Figure 6, and for individual studies - Supplementary Figures S5 and S6). Overall, studies performed before the 1950s did not report on multiple domains, especially participant selection and use of index tests. In newer studies there was a high risk of bias in Domain 1 (participant selection) due to case control designs and variability in reference standards used, with some reference standards less likely to correctly classify true TB status.

**Figure 6:**
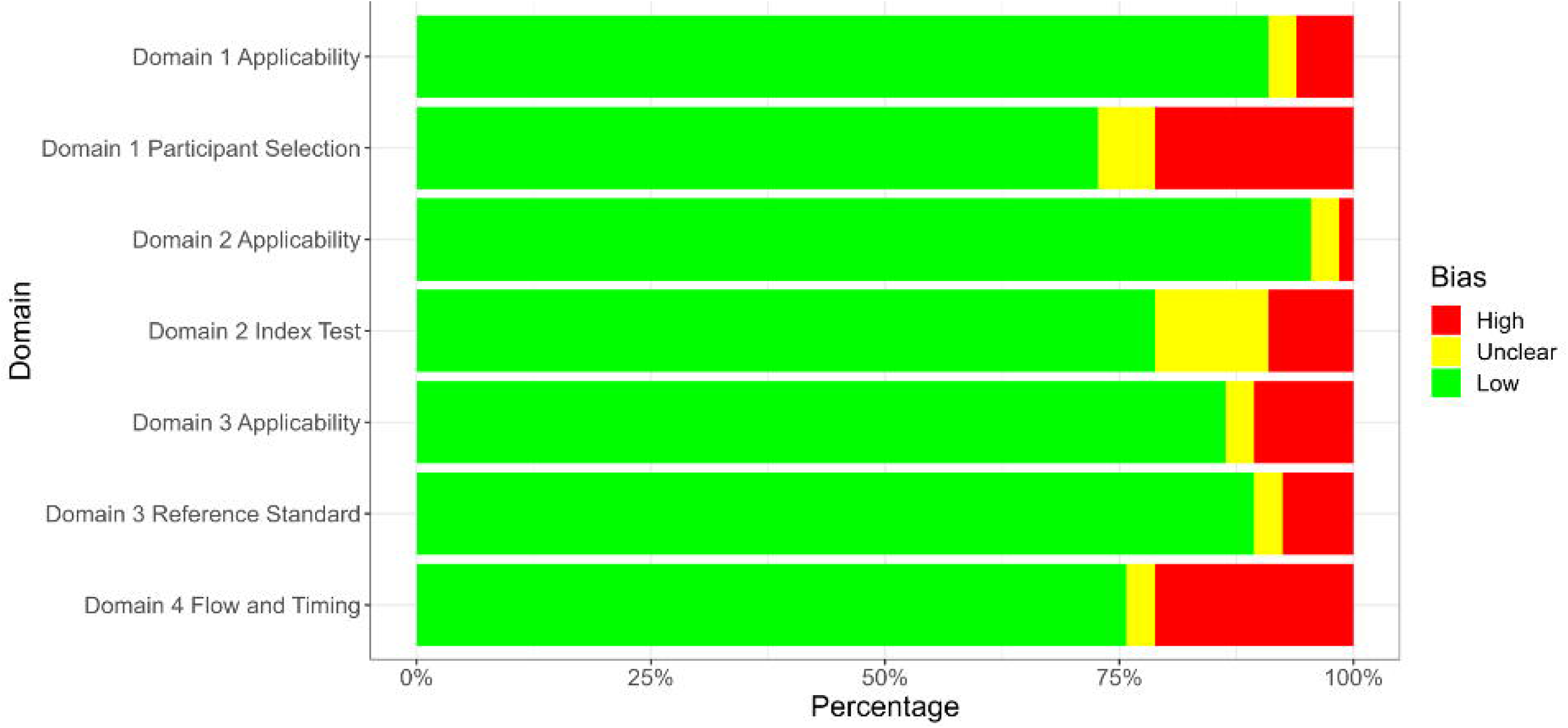
A bar chart to show the percentage of studies with High, Low or unclear bias for each domain.

## Discussion

Diagnosing PTB is challenging, and new, accurate sampling approaches that utilise easier-to-obtain specimens are urgently required. In this systematic review and meta-analysis, which included data on 24,899 sample comparisons from studies conducted over an 89-year period between 1933 and 2022, we found that three upper respiratory tract sample types show promise for accurately diagnosing PTB: laryngeal swabs with a pooled sensitivity of 57.8% and a specificity of 93.8%; NPA with a pooled sensitivity and specificity of 65.2% and 97.9%; and oral swabs with 56.7% and 91.3% against microbiological reference standards.

These accuracy estimates are similar to currently approved TB diagnostic tools on non-sputum samples such as urine lateral flow lipoarabinomannan assays in those who are HIV-positive (80) and stool in children (3). Studies that used laryngeal swabs as a sample type were mostly from Europe in the early 20^th^ century and gave a pooled sensitivity of 57.8% against microbiological reference standards (gastric lavage or sputum culture), here there was large variation in swabbing and culture methodologies used over time as new medias and techniques were invented, and all were conducted before the advent of molecular diagnostics. The studies were mostly carried out among outpatients being investigated for TB, who were unable to produce sputum, with the predominant reference standard being culture of gastric lavage. Use of laryngeal swab testing was discontinued with the advent of chemotherapy and the rapid decline of TB cases in Northern Europe and the closure of sanatoria (81,82), however the original rationale for investigating this sample type, needing an outpatient test that was quick and well-tolerated, reflects similar diagnostic challenges still being faced in high burden countries and the requirements of the WHO TPP (4). Laryngeal swabs may hold promise as an alternative to expand access to TB diagnosis if re-evaluated in prospective studies using modern diagnostic platforms and culture.

The second sample type reviewed was NPA, which were mostly studied in children. NPAs had a sensitivity 65.5% compared to culture of gastric aspirate or induced sputum samples. However as microbiological tests of TB in children have a low sensitivity(83) we also compared NPA to a clinical reference standard for TB, where sensitivity fell to 9%. This reflects the very high proportion of children who were clinically diagnosed with TB in the absence of any positive microbiological test: a common occurrence in clinical practice which reflects the urgent need for improved diagnostic strategies in children. NPA and laryngeal swabs have the benefit of being able to be performed as an outpatient test rather than requiring admission, which could make them a viable alternative in settings where access to hospital services and admission are limited due to cost, distance, and availability. Further research is needed to investigate whether sensitivity can be improved by optimising sample preparation, collection or analysis, how NPA performs in adults, and how NPA could be used within diagnostic algorithms to increase microbiological diagnosis (50).

Oral swabs gave a pooled sensitivity of 56.7% against microbiological reference standards. However, many of the studies that evaluated oral swabs were case control studies with small numbers which have a high risk of bias and would be expected to over-estimate sensitivity. There was also a wide variability in the swab types, sampling methodologies, and specimen processing and analysis. Nevertheless, oral swabs do offer an easy option for sample collection and transport, potentially widening access to TB diagnosis. Further research is needed including prospective diagnostic evaluations to find standardised methodologies and assess its accuracy in a wide range of populations, including children.

Limitations for this systematic review and meta-analysis include the variability of the methods of sample collection and analysis. We mitigated against this by stratifying studies into groups of sample types for meta-analysis; however, some within-group variability remains. For some studies from the early 20^th^ century, we were unable to identify the full text despite extensive searches with librarians, and especially if they were published in a language other than English; this may have biased results to studies in more prominent journals that were more likely to be archived in libraries and those in English. Newer studies, particularly those that examined the accuracy of oral swabs, mainly used case control designs, and had small sample sizes.

Although upper respiratory tract sampling seems a promising avenue for improving access to TB diagnostic testing, further prospective studies are needed to optimise sampling and microbiological methodologies to maximise sensitivity. In children, where novel diagnostics are urgently needed NPA, potentially laryngeal swabs, and a combination of sample sites may offer an alternative sampling methodology that can be used in outpatient settings to widen access. Historical methods using laryngeal swabs showed similar sensitivity and higher specificity than modern studies using oral swabbing in much larger numbers of patients, some of whom were unable to expectorate. Updated evaluations of laryngeal swabbing is required to determine whether these findings are replicated in modern diagnostic accuracy evaluations and across a range of populations. Oral swabs are simple to collect and transport, however this review has shown that more prospective study data is needed to understand whether sensitivity is sufficient for use in clinical practice. If so, this may give a sample type that could be used in outpatient settings in children and adults who are unable to expectorate or in an inpatient setting in those too unwell to produce a sample. Overall upper respiratory tract sampling may offer an alternative non-invasive sample type that can be used to increase access to microbiological diagnosis for Mtb.

## Data Availability

Data and code to reproduce analysis are available at: https://osf.io/9nuvq/

## Contributors

The study was conceived by HRS and LEC. The study design was developed by HRS, LEC and PMP. Data extraction was conducted by HRS, MLO, MS and BR. Data analysis and interpretation were conducted by HRS, PMP, HMR and RMB. The initial manuscript was prepared by HRS and PMP. All authors edited and approved the final manuscript (HRS, HMR, RMB, MLO, MS, BR, LEC and PMP).

## Data sharing

Data of studies and analytical code available at: https://osf.io/9nuvq/

## List of tables

Table 1: Study details included studies. List of figures

Supplementary Table S1: Search strategy S2: Excluded studies

S3: Full study information

S4: Table of data from laryngeal swab reports used in meta-analysis.

S5: Table of data from naso-pharyngeal aspirate reports used in meta-analysis.

S6: Table to show definition of confirmed, probable or not PTB by study.

S7: Table of data from naso-pharyngeal aspirate reports and clinical diagnosis as a reference standard used in the meta-analysis.

S8: Data table to show analysis of reports of oral swabs as sample type.

S9: Data table to show analysis of reports of other sample type data. Supplementary Figures:

SF1: Sensitivity and specificity of laryngeal sampling for active pulmonary tuberculosis, with random effects meta-analysis

SF2: Sensitivity and specificity of nasopharyngeal aspirate for active pulmonary tuberculosis, with random effects meta-analysis

SF3: Sensitivity and specificity of nasopharyngeal aspirate for active pulmonary tuberculosis when clinical diagnosis used as a reference standard, with random effects meta-analysis

SF4: Sensitivity and specificity of oral swab for active pulmonary tuberculosis, with random effects meta-analysis

SF5: Bias of individual studies presented via each domain question and overall rating.

SF6: Bias of individual studies presented by domain.

