## Supplementary material for "Accuracy of upper respiratory tract samples to diagnose Mycobacterium tuberculosis: a systematic review and meta-analysis"

Supplementary data tables and figures

S1: Search strategy

S2: Excluded studies

S3: Full study information

S4: Table of data from laryngeal swab reports used in meta-analysis.

S5: Table of data from naso-pharyngeal aspirate reports used in meta-analysis.

S6: Table to show definition of confirmed, probable or not PTB by study.

S7: Table of data from naso-pharyngeal aspirate reports and clinical diagnosis as a reference standard used in the meta-analysis.

S8: Data table to show analysis of reports of oral swabs as sample type.

S9: Data table to show analysis of reports of other sample type data.

Supplementary Figures:

SF1: Sensitivity and specificity of laryngeal sampling for active pulmonary tuberculosis, with random effects meta-analysis

SF2: Sensitivity and specificity of nasopharyngeal aspirate for active pulmonary tuberculosis, with random effects meta-analysis

SF3: Sensitivity and specificity of nasopharyngeal aspirate for active pulmonary tuberculosis when clinical diagnosis used as a reference standard, with random effects meta-analysis

SF4: Sensitivity and specificity of oral swab for active pulmonary tuberculosis, with random effects meta-analysis

SF5: Bias of individual studies presented via each domain question and overall rating.

SF6: Bias of individual studies presented by domain.

Table S1:

**Search strategy for systematic review**

Search terms

(“Mycobacterium tuberculosis” OR “Mycobacterium tuberculosis complex” OR “tuberculosis” OR “TB” OR “consumption” OR “wasting” OR “thysis” OR “pthysis OR “Koch’s Disease”)

AND

(“oral swab” OR “oral” OR “laryngeal swab” OR “laryngeal” OR “tonsils” OR “saliva” OR “Waldemeyers ring” OR “pharynx” OR “pharyngeal”)

AND

(“diagnosis” OR “diagnostics” OR “samples” OR “sampling”)

Plus database specific strategy terms

Medline (1847-1950): Human

Medline (1950 – current): Human, Clinical Key: Diagnosis

Global Health: As above, remove consumption

Global Health archive: As above, remove consumption

Table S2:

**Excluded studies with reasons**

| Title | Year | Journal | First Author | | Exclusion reason |
| --- | --- | --- | --- | --- | --- |
| On Pulmonary Consumption, and on Bronchial and Laryngeal Disease, &c | 1847 | The Medico-chirurgical review | | | Full text unavailable |
| On Pulmonary Consumption; and on Bronchial and Laryngeal Disease; with Remarks on the Places of Residence Chiefly Resorted to by the Consumptive Invalid | 1853 | Edinburgh medical and surgical journal | | | Wrong study type - case report |
| The Pathology and Treatment of Pulmonary Tuberculosis; and on the Local Medication of Pharyngeal and Laryngeal Diseases, Mistaken for, or Associated with, Phthisis | 1854 | Edinburgh medical and surgical journal | | | No relevant testing |
| Contribution to the History of Laryngeal Phthisis | 1875 | Medico-chirurgical transactions | Marcet, W. | | Background article |
| The Laryngeal Complications of Consumption | 1882 | British medical journal | Williamson, J. M. | | No relevant testing |
| Phthisis of the Larynx | 1883 | Edinburgh medical journal | Mackenzie, G. Hunter | | No relevant testing |
| Some Questions with Regard to Tuberculosis of the Upper Air Passages | 1892 | Transactions. Medico-Chirurgical Society of Edinburgh | M'Bride, P. | | Wrong outcome |
| Clinical Remarks ON A CASE OF TUBERCULOUS DISEASE OF THE LUNGS AND LARYNX, SHOWING THE NEED FOR THE COMPULSORY NOTIFICATION OF PHTHISIS | 1906 | British medical journal | Bramwell, B. | | wrong study design |
| Tuberculosis of the Larynx | 1908 | Proceedings of the Royal Society of Medicine | Hill, W. | | Wrong study design - case report |
| The Throat and Nose in the Ã†tiology of Tuberculosis | 1909 | The Hospital | | | Background article |
| Tuberculosis of the Air-Passages above the Larynx | 1910 | The Hospital | | | Wrong outcome |
| Tuberculosis of the Tonsil, associated with Tuberculous Glands of Neck | 1910 | Proceedings of the Royal Society of Medicine | Carmichael, E. S. | | Wrong outcome |
| Tuberculosis of Pharynx | 1910 | Proceedings of the Royal Society of Medicine | Wylie, A. | | wrong study design |
| Chronic Tuberculosis of the Nose, Larynx, and Lungs | 1911 | Proceedings of the Royal Society of Medicine | Parker, C. A. | | wrong study design |
| Laryngeal Tuberculosis | 1912 | Proceedings of the Royal Society of Medicine | Donelan, J. | | wrong study design |
| Microscopical Sections from a Case of Tuberculous Ulcer of the Larynx; the First suggestive of Epithelioma, the Second of Non-bacillary Tuberculosis? Lupus | 1912 | Proceedings of the Royal Society of Medicine | Grant, J. D. | | wrong study design |
| Some Observations on Tuberculosis of the Nose and Pharynx | 1912 | Bristol medico-chirurgical journal (1883) | Wright, A. J. | | Wrong study design - case report |
| A Post-mortem Specimen of Laryngeal Tuberculosis | 1913 | Proceedings of the Royal Society of Medicine | Davis, E. D. | | Wrong study design - case report |
| What Relation, if any, have the Faucial Tonsils to Pulmonary Tuberculosis? | 1913 | Transactions of the American Climatological Association for the year ... American Climatological Association | Ingals, E. F. | | No relevant testing |
| Tuberculosis of Pharynx and Larynx | 1913 | Proceedings of the Royal Society of Medicine | McKenzie, D. | | wrong study design |
| Some of the Problems of Private Sanatoria for Tuberculosis as observed during Ten Years' Experience in the Pottenger Sanatorium for Diseases of the Lungs and Throat | 1914 | Transactions of the American Climatological and Clinical Association. American Climatological and Clinical Association | Pottenger, F. M. | | Background article |
| THREE YEARS' SANATORIUM EXPERIENCE OF LaRYNGEAL TUBERCULOSIS | 1914 | British medical journal | Thomson, S. | | No relevant testing |
| Tuberculosis of the Larynx | 1920 | Transactions of the American Climatological and Clinical Association. American Climatological and Clinical Association | Forster, A. M. | | Background article |
| Two Cases of Pulmonary Tuberculosis with Laryngeal Symptoms | 1923 | Proceedings of the Royal Society of Medicine | Franklin, P. | | wrong study design |
| Treatment of Laryngeal Tuberculosis in Sanatoria | 1923 | Transactions of the American Climatological and Clinical Association. American Climatological and Clinical Association | Parfitt, C. D. | | Wrong outcome |
| Tuberculosis of the Larynx | 1923 | Proceedings of the Royal Society of Medicine | Renshaw, J. A. | | wrong study design |
| TUBERCULOSIS OF THE LARYNX: Sir StClair Thomson's Report | 1924 | British medical journal | | | No relevant testing |
| Tuberculosis of the Larynx-Its Diagnosis and Treatment: A Plea for Closer Co-operation Between the Physician and Laryngologist | 1924 | Canadian Medical Association journal | Pentecost, R. S. | | Background article |
| Tuberculosis of Nasal Bones | 1924 | Proceedings of the Royal Society of Medicine | Ridout, C. A. | | wrong study design |
| The Mitchell Lecture ON TUBERCULOSIS OF THE LARYNX: ITS SIGNIFICANCE TO THE PHYSICIAN: Deliverd before the Royal College of Physicians of London, November 6th, 1924 | 1924 | British medical journal | Thomson, S. | | No relevant testing |
| Tuberculosis of Larynx | 1927 | Proceedings of the Royal Society of Medicine | Layton, T. B. | | wrong study design |
| Infection of the Upper Respiratory Tract as an Etiological Factor in Pulmonary Disease | 1930 | Canadian Medical Association Journal | Smith, I. R. | | Background article |
| Primary Infection and Pulmonary Tuberculosis in Adults. / Om Primaerinfektionen og Lungetuberkulosen hos Voksne | 1931 | Ugeskrift for Laeger | Heckscher, H. | | Full text not available |
| Tuberculosis of Larynx | 1931 | Proceedings of the Royal Society of Medicine | Wylie, A. | | wrong study design |
| INCIDENCE OF TUBERCULOSIS OF THE TONSIL | 1932 | Canadian Medical Association journal | Heaton, T. G. | | Background article |
| Tubercle Bacilli in the Gastric Contents of Tuberculous Children. A Study of 59 Cases | 1934 | American Review of Tuberculosis and Pulmonary Diseases | Gourley, Ina | | Full text not available |
| Present Concepts of Tuberculous Infection and Disease. Their Principles and Application | 1935 | American Review of Tuberculosis and Pulmonary Diseases | Opie, E. L. | | Full text not available |
| Tuberculosis in Some Rare Situations, Namely Tonsils and Uterus | 1936 | The Indian medical gazette | Krishnaswamy, K. G. | | Background article |
| The Importance of Gastric Lavage for the Demonstration of Tubercle Bacilli in Adults | 1937 | Acta Medica Scandinavica | Gullbring, A. | | Wrong sample type |
| Tuberculosis of the Larynx | 1939 | Proceedings of the Royal Society of Medicine | Suggit, S. | | wrong study design |
| The Routine Examination for Tubercle Bacilli in Sputum | 1940 | Tubercle | Hunter, R. A. | | Wrong sample type |
| Gastric Contests in Tuberculous Children | 1941 | American Review of Tuberculosis and Pulmonary Diseases | Floyd, C | | Wrong sample type |
| Observations on Tuberculosis of the Larynx | 1941 | Edinburgh medical journal | Martin, G. Ewart | | Background article |
| Discussion on Primary Tuberculosis in Adolescents and Adults | 1942 | Proceedings of the Royal Society of Medicine | Kayne, G. G | | Background article |
| The Clinical Value of Faeces Examination for Tubercle Bacilli in the Course of Pulmonary Tuberculosis. Results of a New Method of Examination. / Ueber den klinischen Wert des Stuhlnachweises von Tuberkelbazillen im Verlaufe der Lungentuberkulose. Ergebnisse eines neuen Untersuchungsverfahrens | 1942 | Deutsche Medizinische Wochenschrift | Wolf, J. E. | | Wrong sample type |
| Tuberculosis of the Tonsils | 1943 | The American journal of pathology | Rather, L. J. | | Background article |
| The Detection of Tubercle Bacilli by Deep Culture in Kirehner's Fluid Medium. / Der Nachweis der Tuberkelbazillen in der Tiefenkultur im flÃ¼ssigen NÃ¤hrboden nach Kirchner | 1943 | Zentralblatt fur Bakteriologie, Parasitenkunde, Infektionskrankheiten und Hygiene | Sula, L. | | Wrong outcome - laboratory validation |
| Pulmonary Lavage in the Diagnosis of Tuberculosis. / O lavado pulmonar. No diagnÃ³stico etio-patogÃ©nico ou evolutivo da tuberculose | 1944 | Boletin de la Oficina Sanitaria Panamericana | De Abreu, M. | | Wrong outcome |
| Tubercle Bacilli in the Stomach Contents of Healthy, Normal Adults exposed to Tuberculosis | 1944 | American Review of Tuberculosis and Pulmonary Diseases | Smith, C. R. | | Wrong outcome |
| Tonsillectomy for tuberculosis | 1945 | Schweizerische Zeitschrift fur Tuberkulose. Revue suisse de la tuberculose. Rivista svizzera della tubercolosi | Escher, F. | | Full text unavailable |
| About the oral tubercular primary complex and the post-primary lupoid of the gingiva | 1945 | Schweizerische medizinische Wochenschrift | Nager, : Fanconi | | Full text unavailable |
| Tuberculosis of the tonsils | 1946 | Archives of otolaryngology | Bernstein, D. | | Upper Resp TB |
| Laryngeal tuberculosis | 1946 | Diseases of the chest | Humphries, M. K., Jr. | | Full text unavailable |
| Tuberculous ulcer of the tongue | 1946 | Prensa medica argentina | Steinberg, I. R. | | Full text unavailable |
| Tuberculous sinuses | 1946 | The Medical journal of Australia | Stokes, E. H. | | Full text unavailable |
| Pulmonary, laryngeal, and intestinal tuberculosis and pulmonary water cyst | 1947 | Gaceta medica de Lima | | | Full text unavailable |
| LARYNGEAL tuberculosis | 1947 | Manitoba medical review | | | Full text unavailable |
| Tuberculosis of the mouth | 1947 | Annals of dentistry | Brodsky, R. H. | | Full text unavailable |
| On the functional pathology of laryngeal tuberculosis; comparative clinical and histological examinations | 1947 | Archiv fur Ohren-, Nasen- und Kehlkopfheilkunde | Eschweiler, H. | | Full text unavailable |
| [Tuberculosis of the middle ear and larynx] | 1947 | Jornal do medico | Larroude, C. | | Full text unavailable |
| Cutaneous and laryngeal tuberculosis and Charpy method | 1947 | Maroc medical | Lepinay | | Upper Resp TB |
| Oral tuberculosis | 1947 | Revista dental de Chile | Louvel Bert, R. | | Full text unavailable |
| Tuberculous tonsillitis | 1947 | Schweizerische medizinische Wochenschrift | Taillens, J. P. | | Full text unavailable |
| Laryngeal tuberculosis in infancy | 1947 | Policlinico infantile | Tavani, E. | | Full text unavailable |
| Tuberculosis of the oral mucosa | 1948 | Deutsche dentistische Zeitschrift | Bachmann, W. | | Full text unavailable |
| Tuberculous laryngitis; a controlled study | 1948 | American review of tuberculosis | Black, J. P. M. | | Full text unavailable |
| Comments on laryngeal tuberculosis | 1948 | L'union medicale du Canada | Brahy, J. | | Full text unavailable |
| Tuberculosis of the larynx | 1948 | Acta oto-rhino-laryngologica Belgica | De Prest, R. A. | | Full text unavailable |
| On tuberculosis of the nose, pharynx and tonsils | 1948 | Practica medica | Jimenez Encina, C. | | Full text unavailable |
| Contribution to the study of the so-called latent tuberculosis of the lymphatic ring of Waldeyer | 1948 | Practica oto-rhino-laryngologica | Langraf, F. | | Full text unavailable |
| Laryngeal tuberculosis; observations based on an experience of 28 years with laryngeal tuberculosis | 1948 | The Annals of otology, rhinology, and laryngology | Looper, E. A | | Wrong outcome |
| Tuberculosis infection of the palatine tonsils | 1948 | Revista espanola de tuberculosis | Vassallo De Mumbert, A. | | Full text unavailable |
| Tuberculosis of the tongue | 1948 | La Semana medica | Vergelin, H. E | | Full text unavailable |
| Tonsil tuberculosis in childhood | 1949 | Schweizerische medizinische Wochenschrift | Banhidy, F. | | Duplicate |
| Tuberculosis of the Tonsils in Infancy. / Tonsillentuberkulose im Kindesalter | 1949 | Schweizerische Medizinische Wochenschrift | BÃNhidy, F. | | Wrong outcome |
| Tubercle Bacilli in Pathological Material from Patients with Miliary Tuberculosis : Study of 114 Cases. / La recherche du bacille tuberculeux dans les tuberculoses micronodulaires. (Etude sur 114 cas.) | 1949 | Bulletin et Memoires de la Societe Medicale des Hopitaux de Paris | Bernard, E | | Wrong sample type |
| Nasal mucosal tuberculosis | 1949 | Nordisk medicin | Froste, N. | | Full text unavailable |
| Tuberculosis of the Nasal Mucous Membranes. / NÃ¤sslemhinnetuberkulos | 1949 | Nordisk Medicin | Froste, N. | | wrong study design |
| Tuberculous larynx | 1949 | Archivos medicos mexicanos | Fumagallo Perez, L. | | Full text unavailable |
| Tuberculosis of the tongue | 1949 | Dental items of interest | Gergely, L | | Full text unavailable |
| Laryngeal tuberculosis | 1949 | Archives of otolaryngology | Looper, E. A. | | Wrong outcome |
| [Tuberculosis of the larynx] | 1950 | Medicina, cirurgia, farmacia | Blundi, E | | Full text unavailable |
| [Laryngeal tuberculosis] | 1950 | Acta oto-rhino-laryngologica Belgica | Dupont, P. | | Full text unavailable |
| [Statistical data on the frequency and evolution of laryngeal tuberculosis before the use of streptomycin] | 1950 | Journal de medecine de Lyon | Piaget, F | | Wrong outcome |
| Failure in the Demonstration of Tubercle Bacilli in Gastric Washings. / Svikt i pÃ¡visingen av tuberkelbasiller i ventrikkelskyllevann | 1951 | Nordisk Medicin | Rambl, K. | | Wrong sample type |
| A Nonchromogenic Culture of an Acid-Fast Bacillus isolated from the Nasal Mucus of a Leprosy Patient; its Virulence for Laboratory Animals | 1952 | International Journal of Leprosy | De Souza-Araujo, H. C. | | Wrong outcome |
| Tuberculosis in Childhood, as disclosed or confirmed by Cultivation of Mycobacterium tuberculosis from Gastric Content | 1952 | Medical Journal of Australia | Webster, R. | | Wrong sample type |
| Unclassified Mycobacteria in the Gastric Contents of Healthy Personnel and of Patients of a Tuberculosis Hospital | 1960 | American Review of Respiratory Disease | Atwell, R. J | | Wrong sample type |
| Pulmonary tuberculosis in dog diagnosed by a positive culture of Mycobacterium tuberculosis from laryngeal swab | 1967 | American Review of Respiratory Disease | Trujillo-Rojas, R. A. | | No relevant testing |
| LARYNGEAL TUBERCULOSIS | 1977 | American Journal of Roentgenology | Lindell, M. M | | Wrong outcome |
| Diagnosis of nasopharyngeal tuberculosis by detection of tuberculostearic acid in formalin fixed, paraffin wax embedded tissue biopsy specimens | 1988 | Journal of clinical pathology | Arnold, M | | Wrong outcome |
| LARYNGEAL TUBERCULOSIS REVISITED | 1992 | American Family Physician | Riley, E. C. | | Full text unavailable |
| An exceptional localization of upper respiratory tract tuberculosis: The tonsil | 1996 | Semaine Des Hopitaux | Raji, A | | Wrong study design - case report |
| Risk of tuberculosis transmission in dentistry | 1997 | AAOHN Journal | Murphy, D. C | | No relevant testing |
| Search for Mycobacterium paratuberculosis DNA in orofacial granulomatosis and oral Crohn's disease tissue by polymerase chain reaction | 1997 | Gut | Riggio, M. P | | Wrong pathogen |
| Diagnosis of tuberculosis in sputum negative patients in Dar es Salaam | 1999 | East African Medical Journal | Aris, E. A | | Full text unavailable |
| Changing trends in clinical manifestations of laryngeal tuberculosis | 2000 | Laryngoscope | Shin, J. E | | Wrong outcome |
| Laryngeal tuberculosis | 2000 | American Journal of Otolaryngology | Yencha, M. W | | Wrong outcome |
| Laryngeal tuberculosis | 2004 | Lancet Infectious Diseases | Krecicki, T | | wrong study design |
| A comparative study of the diagnosis of pulmonary tuberculosis using conventional tools and polymerase chain reaction | 2006 | Indian Journal of Tuberculosis | Kavita, Modi-Parekh | | Wrong sample type |
| Tuberculosis of tonsil associated with pulmonary foci | 2008 | Indian Journal of Otolaryngology and Head & Neck Surgery | Santosh, U. P | | wrong study design |
| Laryngeal involvement in patients with active pulmonary tuberculosis | 2008 | European Archives of Oto-Rhino-Laryngology | Topak, M | | Upper Resp TB |
| Is routine pathological examination required in South African children undergoing adenotonsillectomy? | 2009 | SAMJ - South African Medical Journal | Lierop, A. C. van | | Background article |
| Laryngeal tuberculosis | 2010 | Journal of Otolaryngology-Head & Neck Surgery | Cherkaoui, A | | wrong study design |
| Clinical utility of a commercial LAM-ELISA assay for TB diagnosis in HIV-infected patients using urine and sputum samples | 2010 | PloS one | Dheda, Keertan | | Wrong outcome |
| 18F-FDG PET/CT findings of pharyngeal tuberculosis | 2010 | Annals of Nuclear Medicine | Ito, K. | | Background article |
| Tuberculosis of the oral cavity: a systematic review | 2010 | European Journal of Oral Sciences | Kakisi, O. K. | | Background article |
| The TDR tuberculosis specimen bank: a resource for diagnostic test developers | 2010 | International Journal of Tuberculosis and Lung Disease | Nathanson, C. M. | | Background article |
| Nasal tuberculosis--an update of current clinical and laboratory investigation | 2011 | The Journal of laryngology and otology | Masterson, L. | | wrong study design |
| Detection and identification of Mycobacterium tuberculosis and Mycobacterium bovis from clinical species using DNA microarrays | 2012 | Journal of Veterinary Diagnostic Investigation | Jia, K | | Wrong population |
| Laryngoscopic characteristics and diagnosis of laryngeal tuberculosis | 2012 | Journal of Otolaryngology and Ophthalmology of Shandong University | Yu, Ping | | wrong study design |
| Assessment of the Xpert MTB/RIF assay for diagnosis of tuberculosis with gastric lavage aspirates in children in sub-Saharan Africa: a prospective descriptive study | 2013 | The Lancet. Infectious diseases | Bates, Matthew | | Wrong sample type |
| Secondary laryngeal tuberculosis revisited | 2015 | Lung India | Lodha, J. V | | wrong study design |
| Morphological aspects in tuberculosis of oral cavity our experience and a review of the literature attempt | 2015 | Romanian Journal of Morphology and Embryology | Popescu, M. R | | wrong study design |
| Tuberculous cheilitis revealing pulmonary tuberculosis. / ChÃ©ilite tuberculeuse rÃ©vÃ©lant une tuberculose pulmonaire | 2016 | Pan African Medical Journal | Bricha, M | | Upper Resp TB |
| Tuberculous cheilitis revealing pulmonary tuberculosis | 2016 | Pan African Medical Journal | Bricha, M | | Duplicate |
| Respiratory microbes present in the nasopharynx of children hospitalised with suspected pulmonary tuberculosis in Cape Town, South Africa | 2016 | BMC Infectious Diseases | Dube, Felix S | | No relevant testing |
| Nasal swab real-time PCR is not suitable for in vivo diagnosis of bovine tuberculosis | 2017 | Pesquisa VeterinÃ¡ria Brasileira | Mayer, F. Q | | Wrong population |
| ORAL LOCALIZATION OF TUBERCULOSIS: CASE REPORT AND LITERATURE REVIEW | 2017 | Journal of Medical and Surgical Research | Nabih, O | | wrong study design |
| Performance of the Xpert MTB/RIF assay in the diagnosis of tuberculosis in formalin-fixed, paraffin-embedded tissues | 2017 | International journal of mycobacteriology | Polepole, Pascal | | Wrong sample type |
| Repertoire of bacterial species cultured from the human oral cavity and respiratory tract | 2018 | Future Microbiology | Fonkou, M. D. M | | wrong study design |
| GeneXpert MTB/RIF Outperforms Mycobacterial Culture in Detecting Mycobacterium tuberculosis from Salivary Sputum | 2018 | BioMed Research International | Shi, Jin | | Wrong sample type |
| Detection of Mycobacterium tuberculosis purified ESAT-6 (Rv3875) by magnetic bead-coupled gold nanoparticle-based immuno-PCR assay | 2018 | International Journal of Nanomedicine | Singh, N | | Wrong outcome - lab validation |
| Laryngeal tuberculosis diagnosed in a pathological laboratory in Senegal (2011-2015). / La tuberculose laryngÃ©e diagnostiquÃ©e dans un laboratoire d'anatomie pathologique du sÃ©nÃ©gal (2011-2015) | 2018 | Bulletin de la SociÃ©tÃ© de Pathologie Exotique | Thiam, I | | wrong study design |
| Molecular Detection of Mycobacterium tuberculosis from Stools in Young Children by Use of a Novel Centrifugation-Free Processing Method | 2018 | Journal of clinical microbiology | Walters, Elisabetta | | Wrong sample type |
| Guidance for Studies Evaluating the Accuracy of Biomarker-Based Nonsputum Tests to Diagnose Tuberculosis | 2019 | The Journal of infectious diseases | Drain, Paul K | | Background article |
| Lipoarabinomannan in sputum to detect bacterial load and treatment response in patients with pulmonary tuberculosis: Analytic validation and evaluation in two cohorts | 2019 | PLoS medicine | Kawasaki, Masanori | | Wrong sample type |
| Urine Xpert MTB/RIF for the diagnosis of childhood tuberculosis | 2019 | International Journal of Infectious Diseases | Lopez, A. L | | Wrong outcome |
| Liquid mycobacterial culture outcomes after different sputum collection techniques before and during treatment | 2019 | Tuberculosis (Edinburgh, Scotland) | Lourens, Madeleine | | Wrong sample type |
| Prevalence of oral lesions in tuberculosis: a cross sectional study | 2019 | Journal of Family Medicine and Primary Care | Purnendu, Rout | | wrong study design |
| Sample adequacy controls for infectious disease diagnosis by oral swabbing | 2020 | PloS one | Deviaene, Meagan | | Wrong outcome - lab validation |
| Tonsillar Tuberculosis | 1945 | Fichero medico terapeutico | Hamuy, D. J | | Full text unavailable |
| [Examination of stomach contents and laryngeal mucus in adult tubercular patients] | 1945 | Problemy tuberkuleza | Klebanova, A. A. | | Full text unavailable |
| Laryngeal tuberculosis | 1946 | Archives of otolaryngology | Auerbach, O. | | Background article |
| Tonsils and tuberculosis | 1946 | Medecine et hygiene | Vetter, H. | | Full text unavailable |
| Subcutaneous gum tuberculosis and pulmonary tuberculosis | 1947 | Le Poumon | Delord, M. | | Full text unavailable |
| Tonsils and tuberculosis | 1947 | Schweizerische medizinische Wochenschrift | Vetter, H. | | Full text unavailable |
| About the development of tonsil tuberculosis | 1947 | Schweizerische Zeitschrift fur Pathologie und Bakteriologie. Revue suisse de pathologie et de bacteriologie | Wegelin, C. | | Full text unavailable |
| Histological research on the participation of tonsillary lymphatic tissue in the tubercular process | 1948 | Annali dell'Istituto "Carlo Forlanini" | Colantuono, P. | | Full text unavailable |
| Tomography of larynx in disseminated pulmonary tuberculosis | 1949 | Diseases of the chest | Espinoza Galarza, M. | | Full text unavailable |
| Hypopharyngeal (Laryngeal) Swabbing for the Cultural Diagnosis of Pulmonary Tuberculosis. A Statement of the Laboratory Subcommittee | 1956 | American Review of Tuberculosis and Pulmonary Diseases | A. J. O'Hea. | | Background article |
| Catarrhal Affections of the Nasal Passages as a Cause of Pulmonary Phthisis, with Special Reference to the Question of Heredity | 1886 | Transactions of the ... Annual Meeting of the American Climatological Association. American Climatological Association. Annual Meeting | Jarvis, W. C. | | Wrong outcome |
| The Tonsil in Tuberculosis | 1898 | The Hospital | | | Wrong test type |
| The Recognition of Early Changes in the Larynx in Tuberculosis | 1913 | Transactions of the American Climatological Association for the year ... American Climatological Association | Casselberry, W. E. | | Wrong test type |
| Laryngeal Lesion associated with Apparent Miliary Tuberculosis of the Lung | 1927 | Proceedings of the Royal Society of Medicine | Howarth, W. | | Wrong study design - case report |
| TUBERCULOSIS OF THE LARYNX | 1929 | British medical journal | Howarth, W. G. | | Background article |
| TUBERCULOSIS OF THE LARYNX | 1929 | British medical journal | Thomson, S. | | Background article |
| Diagnostic Value of Direct and Cultural Examination of Laryngeal Secretion in Tuberculosis. / Importanza diagnostica della ricerca dei bacilli di Koch mediante l'esame diretto e culturale del secreto prelevato in laringe | 1936 | Rivista di Patologia e Clinica della Tuberculosi | Bernabo-Silorata, A | | Full text unavailable |
| Negative Sputum and Pulmonary Tuberculosis | 1937 | Tubercle | Davies, G. I. | | wrong study design |
| The Value of Exact Sputum Examination (Laryngeal Swab Culture) in the Diagnosis and Management of Pulmonary Tuberculosis | 1943 | Tubercle | Munro-Ashman, D | | Wrong study design - case report |
| The incidence, treatment and prognosis of tuberculous laryngitis in pulmonary tuberculosis | 1947 | The Medical press | Lambert, V. | | Full text unavailable |
| Relationship between pulmonary tuberculosis and tonsils | 1948 | Minerva medica | Puricelli, P. J. | | Full text unavailable |
| Contribution to the study of so-called latent tuberculosis of the Waldeyer's lymph ring | 1949 | Schweizerische medizinische Wochenschrift | Langraf, F. | | Full text unavailable |
| Tuberculous laryngitis and tracheo bronchitis | 1949 | The Annals of otology, rhinology, and laryngology | O'Keefe, J. J. | | Full text unavailable |
| Cervical lymph node tuberculosis and the tonsils | 1949 | Acta pathologica et microbiologica Scandinavica | Pentti, M. | | Background article |
| Tonsillar swab culture in lung tuberculosis | 1950 | Acta tuberculosea Scandinavica | Adler, H | | Full text unavailable |
| [Role of the nasopharynx complex in primary tuberculosis] | 1950 | Revue de la tuberculose | Couve, P | | Full text unavailable |
| [Comparative bacteriologic research on secretion withdrawn from the larynx in pulmonary tuberculosis] | 1950 | Archivio italiano di otologia, rinologia e laringologia | Redoglia, F. | | Full text unavailable |
| An investigation of the occurrence of fungi in 250 laryngeal swabs from tuberculous patients | 1953 | Acta Path. et Microb. Scandinavica | Reiersol, S. | | Wrong outcome |
| Direct Trachal Lavage for Rapid Recovery of Mycobacterium tuberculosis from the Respiratory Tract | 1956 | Brit. J. Tuberculosis | Jones, J. S. | | Wrong outcome |
| The Laryngeal Swab Method of diagnosing Pulmonary Tuberculosis | 1956 | Monthly Bull. Ministry of Health & Pub. Health Lab. Service (directed by Med. Res. Council) | Thomas, C. H. H. | | Wrong test type |
| Isolation of mycobacteria from tonsils, naso-pharyngeal secretions and lymph nodes in East Anglia | 1970 | Tubercle | Stewart C, J | | Epidemiological study |
| Sensitivity and specificity of PCR for detection of Mycobacterium tuberculosis: a blind comparison study among seven laboratories | 1994 | Journal of clinical microbiology | Noordhoek, G. T | | Wrong outcome - lab validation |
| Identification of novel tuberculosis diagnostic biomarkers in plasma and saliva | 2016 | European Respiratory Journal | Jacobs, R | | Wrong test type biomarkers |
| Suitability of saliva for Tuberculosis diagnosis: comparing with serum | 2017 | BMC Infectious Diseases | Namuganga, Anna Ritah | | Wrong outcome - lab validation |
| An optimised saliva collection method to produce high-yield, high-quality RNA for translational research | 2020 | Plos One | Sullivan, R | | Wrong outcome |
| Primary tuberculosis cutis orificialis; a different face of the same coin | 2021 | Idcases | Ali, G. A. | | Wrong study type - case report |
| Next-Generation Digital Biomarkers for Tuberculosis and Antibiotic Stewardship: Perspective on Novel Molecular Digital Biomarkers in Sweat, Saliva, and Exhaled Breath | 2021 | Journal of medical Internet research | Brasier, Noe | | Biomarker study - wrong test not micro |
| Primary tonsillar tuberculosis | 2021 | Klimik Dergisi | DaÅŸli, S | | Wrong study type - case report |
| Tuberculosis in the head and neck: changing trends and age-related patterns | 2021 | Laryngoscope | Gehrke, T. | | Wrong study type - case report |
| New developments in tuberculosis diagnosis and treatment | 2022 | Breathe | Gil, C. M. | | Wrong study type - review |
| Oro-facial tuberculosis - Is it still an enigmatic entity? | 2021 | Indian Journal of Pathology and Microbiology | Gupta, L. | | Wrong study type - case report |
| A rare association of tonsillar tuberculosis and lichen scrofulosorum | 2021 | Egyptian Journal of Otolaryngology | Harit, A. | | Wrong study type - case report |
| Dysphagia as the Presenting Symptom of Laryngeal Tuberculosis | 2021 | Cureus | Kandah, E. | | Wrong study type - case report |
| Host-Based Biomarkers in Saliva for the Diagnosis of Pulmonary Tuberculosis in Children: A Mini-Review | 2021 | Frontiers in Pediatrics | Khambati, N | | Biomarker study - wrong test not micro |
| A narrative review of exploring potential salivary biomarkers in respiratory diseases: still on its way | 2021 | Journal of Thoracic Disease | Li, C. X. | | Biomarker study - wrong test not micro |
| Cervical Tuberculosis Combined With Papillary Thyroid Carcinoma With Lateral Neck Metastasis | | Ent-Ear Nose & Throat Journal | Lin, H. Y. | | Wrong study type - case report |
| The history of tuberculosis of the larynx | 2021 | Laryngo-Rhino-Otologie | Luckhaupt, H. | | Wrong study type - review |
| Suspected IgG4-related disease of the submandibular salivary gland and periorbital soft tissue in a man with latent tuberculosis | 2021 | Bmj Case Reports | McGreal-Bellone, A. | | Wrong study type - case report |
| Primary tonsillar tuberculosis in a pediatric patient case report and literature review | 2021 | Medicine (Baltimore) | Moisa, S. M. | | Wrong study type - case report |
| Identification of novel salivary candidate protein biomarkers for tuberculosis diagnosis: a preliminary biomarker discovery study | 2021 | Tuberculosis | Mutavhatsindi, H. | | Biomarker study - wrong test not micro |
| Determining the prevalence of rifampicin resistant tuberculosis in a tertiary care centre of north India by using rapid culture method and gene Xpert | 2021 | Journal of Pure and Applied Microbiology | Pokhriyal, B. C | | Wrong sample type - not upper respiratory tract |
| Laryngeal tuberculosis: a neglected diagnosis | 2022 | Bmj Case Reports | Raj, R. | | Wrong study type - case report |
| Lingual primary tuberculosis mimicking malignancy | 2021 | Annals of Medicine and Surgery | Razem, B. | | Wrong study type - case report |
| Primary tuberculosis of the pyriform sinus: A case report | 2022 | Annals of Medicine and Surgery | Touihmi, S. | | Wrong study type - case report |
| An Analysis of Xpert Test for Diagnosing Maxillofacial Tuberculosis |  | Journal of Maxillofacial & Oral Surgery | Tripathi, R. | | Wrong outcome - diagnosing URT TB |
| A look inside: oral sampling for detection of non-oral infectious diseases | 2021 | Journal of Clinical Microbiology | Valinetz, E. D. | | Wrong study type - review |
| Life-Threatening Stridor due to Laryngeal Tuberculosis in the COVID-19 Era: Report of a Case |  | Ent-Ear Nose & Throat Journal | Valjarevic, S. | | Wrong study type - case report |
| Two cases of tuberculous retropharyngeal abscess in adults | 2021 | The Journal of international medical research | Xu, Xiaofeng | | Wrong study type - case report |
| Case Report: Lingual Tuberculosis Reveaed by a Cold Abcess |  | Indian Journal of Otolaryngology and Head & Neck Surgery | Younes, H | | Wrong study type - case report |
| Laryngeal and voice disorders in patients with pulmonary tuberculosis | 2021 | Iranian Journal of Otorhinolaryngology | Youssef, G | | Wrong outcome - diagnosing URT TB |
| The participation of tonsils in childhood tuberculosis | 1947 | Revista chilena de pediatria | Pena Cereceda, J | | Full text unavailable |
| A Note on the Laryngeal Mirror Test for the Detection of Tubercle Bacilli | 1936 | Tubercle | Wood, W. B. | | Wrong study design - case report |
| Tuberculosis, of Tonsils: Its Relation in Children with Tubercle Bacilli in Gastric Contents | 1941 | American Review of Tuberculosis and Pulmonary Diseases | Rosencrantz, E. | | Wrong test type histology |
| On the occurrence and treatment of laryngeal tuberculosis in patients at the pulmonary clinic | 1945 | Duodecim; laaketieteellinen aikakauskirja | Santalarti, V. | | Background article |
| [Gastric lavage and laryngeal swab in search of Koch's bacillus] | 1949 | Revista brasileira de medicina | Silva Junior, E. | | Review article |
| [Laryngeal tuberculosis associated with pulmonary tuberculosis; its incidence, prognosis and treatment] | 1950 | The Medical press | | | Wrong outcome - URT TB |
| Proteomics in Biomarker Discovery for Tuberculosis: Current Status and Future Perspectives | 2022 | FRONTIERS IN MICROBIOLOGY | | Guo, JB | Wrong test type - biomarkers |
| Improved Conventional and New Approaches in the Diagnosis of Tuberculosis | 2022 | FRONTIERS IN MICROBIOLOGY | | Dong, BY | Review article |
| Performance of Xpert Ultra nasopharyngeal swab for identification of tuberculosis deaths in northern Tanzania | 2022 | Clinical microbiology and Infection | | Costales, C. | Wrong population – post-mortem |

S Table S3:

Full descriptive data including methods used available as excel spreadsheet.

https://osf.io/9nuvq/?view_only=a00f5d373c27461c8b19517673c383aa

S Table S4:

Full data table to show included reports and analysis of laryngeal swabs as sample type

| Study | Comparison | TP | FN | FP | TN | Sens | Spec | Sens_Percentage | Spec_Percentage | LCI_sens | UCI_sens | LCI_spec | UCI_spec |
| --- | --- | --- | --- | --- | --- | --- | --- | --- | --- | --- | --- | --- | --- |
| 1941_Nassau_Discharged 1940 | Sputum culture | 28 | 35 | 35 | 68 | 0.444444 | 0.660194 | 44.44444 | 66.01942 | 0.321741 | 0.567148 | 0.568722 | 0.751666 |
| 1941_Nassau_Inpatient 1940 | Sputum culture | 20 | 18 | 38 | 31 | 0.526316 | 0.449275 | 52.63158 | 44.92754 | 0.367559 | 0.685072 | 0.331906 | 0.566645 |
| 1948_Forbes_Inpatient | Gastric lavage | 22 | 20 | 10 | 48 | 0.52381 | 0.827586 | 52.38095 | 82.75862 | 0.372764 | 0.674855 | 0.730371 | 0.924802 |
| 1948_Forbes_Outpatient | Gastric lavage | 5 | 6 | 8 | 82 | 0.454545 | 0.911111 | 45.45455 | 91.11111 | 0.160288 | 0.748803 | 0.852316 | 0.969907 |
| 1948_Forbes_Mass radiography | Sputum culture | 13 | 15 | 3 | 65 | 0.464286 | 0.955882 | 46.42857 | 95.58824 | 0.279556 | 0.649015 | 0.907072 | 1 |
| 1948_Hounslow_Hospital patients | Gastric lavage | 18 | 14 | 21 | 140 | 0.5625 | 0.869565 | 56.25 | 86.95652 | 0.390618 | 0.734382 | 0.817543 | 0.921588 |
| 1950_Duggan_Inpatients and Outpatients both adults and children | Gastric lavage | 27 | 3 | 5 | 65 | 0.9 | 0.928571 | 90 | 92.85714 | 0.792646 | 1 | 0.868239 | 0.988904 |
| 1950_Renoux_Sanatorium patients | Gastric lavage | 19 | 4 | 4 | 29 | 0.826087 | 0.878788 | 82.6087 | 87.87879 | 0.67118 | 0.980994 | 0.767432 | 0.990144 |
| 1951_Armstrong_Hospital patients | Gastric lavage | 84 | 15 | 105 | 763 | 0.848485 | 0.879032 | 84.84848 | 87.90323 | 0.777855 | 0.919115 | 0.857339 | 0.900726 |
| 1953_Chaves_Clinic patients | Gastric lavage | 115 | 72 | 20 | 1211 | 0.614973 | 0.983753 | 61.49733 | 98.3753 | 0.545229 | 0.684718 | 0.976691 | 0.990816 |
| 1954_Frostad_TB hospital patients | Gastric lavage | 91 | 104 | 88 | 1217 | 0.466667 | 0.932567 | 46.66667 | 93.2567 | 0.396644 | 0.53669 | 0.918961 | 0.946173 |
| 1955_Lind_Patients with lesions | Gastric lavage | 11 | 13 | 2 | 95 | 0.458333 | 0.979381 | 45.83333 | 97.93814 | 0.258987 | 0.657679 | 0.951102 | 1 |
| 1955_Lind_Patients with lesions | Sputum culture | 26 | 56 | 8 | 293 | 0.317073 | 0.973422 | 31.70732 | 97.34219 | 0.216353 | 0.417793 | 0.955251 | 0.991593 |
| 1955_Wallace_Inpatients | Sputum culture | 37 | 14 | 13 | 73 | 0.72549 | 0.848837 | 72.54902 | 84.88372 | 0.60301 | 0.84797 | 0.773129 | 0.924545 |
| 1956_Engbaek_Outpatients | Gastric lavage | 38 | 76 | 10 | 1107 | 0.333333 | 0.991047 | 33.33333 | 99.10474 | 0.246797 | 0.419869 | 0.985523 | 0.996571 |
| 1955_Tonge_Outpatients | Gastric lavage | 52 | 78 | 2 | 333 | 0.4 | 0.99403 | 40 | 99.40299 | 0.315785 | 0.484215 | 0.98578 | 1 |
| 1962_Hsing_Hospital patients | Gastric lavage | 264 | 90 | 46 | 907 | 0.745763 | 0.951731 | 74.57627 | 95.17314 | 0.700403 | 0.791123 | 0.938123 | 0.96534 |
| 1966_Pechacek_TB hospital patients | Homogenised 24 hours sputum | 24 | 39 | 0 | 235 | 0.380952 | 1 | 38.09524 | 100 | 0.261035 | 0.50087 | 1 | 1 |
| 1966_Pechacek_TB hospital patients | Homogenised 24 hours sputum | 28 | 29 | 0 | 170 | 0.491228 | 1 | 49.12281 | 100 | 0.361444 | 0.621012 | 1 | 1 |
| 1968_Lloyd, A. V. C._Inpatients | Gastric lavage | 12 | 5 | 26 | 17 | 0.705882 | 0.395349 | 70.58824 | 39.53488 | 0.489282 | 0.922482 | 0.24921 | 0.541487 |
| 1950_Lundar_Inpatients | Gastric lavage | 38 | 34 | 16 | 216 | 0.527778 | 0.931034 | 52.77778 | 93.10345 | 0.412462 | 0.643094 | 0.898427 | 0.963641 |
| 1952_Roald_Inpatients | Gastric lavage | 96 | 37 | 52 | 298 | 0.721805 | 0.851429 | 72.18045 | 85.14286 | 0.645647 | 0.797962 | 0.814167 | 0.88869 |
| 1952_Smedsrud_Inpatients | Sputum culture | 34 | 21 | 2 | 14 | 0.618182 | 0.875 | 61.81818 | 87.5 | 0.489783 | 0.746581 | 0.712948 | 1 |

Supplementary table S5:

Data table to show analysis of reports of NPA as sample type

| Study | Comparison | TP | FN | FP | TN | Sens | Spec | Sens_Percentage | Spec_Percentage | LCI_sens | UCI_sens | LCI_spec | UCI_spec |
| --- | --- | --- | --- | --- | --- | --- | --- | --- | --- | --- | --- | --- | --- |
| 1998_Franchi_Culture | Gastric aspirate | 17 | 7 | 2 | 38 | 0.708333 | 0.95 | 70.83333 | 95 | 0.526484 | 0.890183 | 0.882458 | 1 |
| 1998_Franchi_PCR | Gastric aspirate | 13 | 4 | 5 | 42 | 0.764706 | 0.893617 | 76.47059 | 89.3617 | 0.563062 | 0.966349 | 0.805468 | 0.981766 |
| 2007_Owens_Culture | Induced sputum | 16 | 3 | 5 | 64 | 0.842105 | 0.927536 | 84.21053 | 92.75362 | 0.678142 | 1 | 0.866364 | 0.988709 |
| 2012_Zar_Culture | Induced sputum | 61 | 23 | 0 | 451 | 0.72619 | 1 | 72.61905 | 100 | 0.630831 | 0.82155 | 1 | 1 |
| 2013_Zar_Xpert MTB/RIF | Induced sputum | 9 | 21 | 8 | 346 | 0.3 | 0.977401 | 30 | 97.74011 | 0.136015 | 0.463985 | 0.961919 | 0.992883 |
| 2019_Hanrahan_Xpert MTB/RIF | Induced sputum | 0 | 2 | 2 | 90 | 0 | 0.978261 | 0 | 97.82609 | 0 | 0 | 0.948461 | 1 |
| 2019_Hanrahan_Culture | Induced sputum | 0 | 0 | 1 | 93 | NA | 0.989362 | NA | 98.93617 | NA | NA | 0.968622 | 1 |
| 2019_Zar_Ultra | Induced sputum | 21 | 19 | 5 | 150 | 0.525 | 0.967742 | 52.5 | 96.77419 | 0.370242 | 0.679758 | 0.939926 | 0.995558 |
| 2021_Song_Xpert MTB/RIF | Gastric aspirate MGIT | 14 | 8 | 3 | 269 | 0.636364 | 0.988971 | 63.63636 | 98.89706 | 0.435347 | 0.83738 | 0.976559 | 1 |
| 2021_Song_MGIT | Gastric aspirate MGIT | 18 | 4 | 4 | 268 | 0.818182 | 0.985294 | 81.81818 | 98.52941 | 0.65701 | 0.979353 | 0.970989 | 0.9996 |

Supplementary table S6:

Table to show definition of confirmed, probable or not PTB by study.

| Paper | Confirmed PTB | Number | Probable PTB | Number | Not TB | Number |
| --- | --- | --- | --- | --- | --- | --- |
| Owens 2007 | presence of a  positive smear or culture for MTB – taken from the probable TB group |  | Probable PTB was suggested by hilar or mediastinal lymphadenopathy, local collapse/consolidation, severe bilateral but  asymmetric disease, cavitation or miliary changes. Chest radiographs were reviewed by JBSC who was blinded to the clinical details. | 94 | No CXR changes as for probable TB not tested | 0 |
| Zar 2012 | Any specimen positive for M. tuberculosis on culture | 87 | All others | 255 | Documented resolution of symptoms and signs at  follow-up in children who didn’t receive tuberculosis treatment | 193 |
| Zar 2013 | Any induced sputum culture  positive for M tuberculosis | 30 | All other children | 167 | Culture negative and documented resolution of symptoms and signs at a follow-up visit at 3 months in children who did not receive tuberculosis treatment | 187 |
| Cakir 2018 |  | 0 | Diagnosis of TB disease was established in accordance with the World Health Organization’s TB Standards  case definition – respiratory symptoms, radiological findings, tuberculin skin test positivity, history of contact with active TB, and acid fast bacillus and MTB culture positivity | 40 |  | 0 |
| Zar 2019 | Culture positive for Mycobacterium tuberculosis (excluding Ultra) | 40 | Culture negative, clinically diagnosed with TB | 104 | culture negative, not clinically diagnosed with TB, no tuberculosis treatment given, and documented improvement at 3-mo follow-up visit | 51 |
| Hanrahan 2018 | Microbiologically using smear, culture or Xpert on any one of the samples collected | 4 | No microbiological confirmation on any sample and at least two of the following: CXR consistent with TB, positive clinic response to  anti-TB treatment, documented exposure to TB or a positive TST. | 100 | Unlikely TB were those  Without mycobacteriological confirmation who did  not meet the criteria for unconfirmed TB. | 15 |
| Osorio 2020 |  |  | (i) signs/symptoms: (a) persistent cough (>2 weeks), unremitting cough; (b) weight loss/failure to thrive; © persistent (>1 week), unexplained fever reported by guardian; (d) persistent, unexplained lethargy or reduced playfulness; (e) infants 0–60 days with additional signs and symptoms like neonatal pneumonia, unexplained hepatosplenomegaly or sepsis-like illness; (ii) findings on chest X-ray congruent with pulmonary TB (presence of lymphadenopathy and/or abnormalities consistent with TB as new infiltrates) and read by two blinded operators (clinician and TB expert); (iii) history of exposure to M. tuberculosis within the preceding 12 months; or (iv) response to antituberculosis treatment yet no acid-fast bacillus on the sputum smear or a negative Xpert MTB/RIF test. | 17 | No TB | 28 |
| Song 2021 |  |  | Participants had to have a visible cervical lymph node mass measuring >1 cm × 1 cm and persisting for >1 month despite antibiotic therapy for at least 5 days or a parenchymal abnormality on chest radiograph in addition to at least one of the following symptoms: 1) cough or wheezing >4 weeks not resolved after treatment with antibiotics, with cough continuing for at least 2 weeks after starting antibiotics (for hospitalized children only, respiratory distress or diagnosis of severe pneumonia not responding to antibiotics after 5 days or any cough >4 weeks despite at least 5 days of antibiotics), 2) moderate or severe malnutrition (defined as weight-for-height Z score <-2 standard deviations (SD) or -3SD, respectively) not responding after 3 weeks of treatment for malnutrition, and 3) reported fever >7 days not responding after 5 days of antibiotics or antimalarials. Children were excluded from the study if they were currently on tuberculosis treatment or isoniazid preventive therapy (IPT) or had received treatment for tuberculosis in the past year or IPT in the last 6 months. | 294 |  |  |

Supplementary table S7:

Data table to show analysis of reports of NPA as sample type using clinical diagnosis as a reference test:

| Study | Clinical_comparison | Probable.PTB | True.positive.PTB | Definite.PTB | True.positive.DTB | No.TB | Combined.True.positive | FN | FP | TN | Sens | Spec | Sens_Percentage | Spec_Percentage | SE_sens | LCI_sens | UCI_sens | SE_spec | LCI_spec | UCI_spec |
| --- | --- | --- | --- | --- | --- | --- | --- | --- | --- | --- | --- | --- | --- | --- | --- | --- | --- | --- | --- | --- |
| 2007_Owens_Culture | Probable PTB | 94 | 21 | 0 | 0 | 0 | 21 | 73 | 0 | 0 | 0.223404 | NA | 22.34043 | NA | 0.042961 | 0.1392 | 0.307609 | NA | NA | NA |
| 2018_Cakir_Culture | Probable PTB | 40 | 5 | 0 | 0 | 0 | 5 | 35 | 0 | 0 | 0.125 | NA | 12.5 | NA | 0.052291 | 0.022509 | 0.227491 | NA | NA | NA |
| 2012_Zar_Culture | Probable and definite = positive, not = negative. | 255 | 0 | 87 | 61 | 193 | 61 | 281 | 0 | 193 | 0.178363 | 1 | 17.83626 | 100 | 0.0207 | 0.13779 | 0.218935 | 0 | 1 | 1 |
| 2012_Zar_Xpert MTB/RIF | Probable and definite = positive, not = negative. | 255 | 7 | 87 | 49 | 193 | 56 | 286 | 1 | 192 | 0.163743 | 0.994819 | 16.37427 | 99.48187 | 0.02001 | 0.124524 | 0.202961 | 0.005168 | 0.98469 | 1 |
| 2013_Zar_Xpert MTB/RIF | Probable and definite = positive, not = negative. | 167 | 5 | 30 | 12 | 187 | 17 | 180 | 0 | 187 | 0.086294 | 1 | 8.629442 | 100 | 0.020006 | 0.047083 | 0.125506 | 0 | 1 | 1 |
| 2019_Hanrahan_Xpert MTB/RIF | Confirmed, unconfirmed, not | 100 | 0 | 4 | 2 | 15 | 2 | 102 | 0 | 15 | 0.019231 | 1 | 1.923077 | 100 | 0.013467 | -0.00716 | 0.045626 | 0 | 1 | 1 |
| 2019_Hanrahan_Culture | Confirmed, unconfirmed, not | 100 | 0 | 4 | 1 | 15 | 1 | 103 | 0 | 15 | 0.009615 | 1 | 0.961538 | 100 | 0.009569 | -0.00914 | 0.028371 | 0 | 1 | 1 |
| 2019_Zar_Ultra | Confirmed, unconfirmed, not | 104 | 5 | 40 | 21 | 51 | 26 | 118 | 0 | 51 | 0.180556 | 1 | 18.05556 | 100 | 0.032054 | 0.117729 | 0.243382 | 0 | 1 | 1 |
| 2021_Song_Xpert MTB/RIF | Clinically suspected | 294 | 17 | 0 | 0 | 0 | 17 | 277 | 0 | 0 | 0.057823 | NA | 5.782313 | NA | 0.013613 | 0.031142 | 0.084504 | NA | NA | NA |
| 2021_Song_MGIT | Clinically suspected | 294 | 22 | 0 | 0 | 0 | 22 | 272 | 0 | 0 | 0.07483 | NA | 7.482993 | NA | 0.015345 | 0.044753 | 0.104907 | NA | NA | NA |
| 2020_Osorio_Xpert MTB/RIF | Clinically suspected | 17 | 0 | 0 | 0 | 28 | 0 | 17 | 0 | 28 | 0 | 1 | 0 | 100 | 0 | 0 | 0 | 0 | 1 | 1 |
| 2020_Osorio_MGIT | Clinically suspected | 17 | 4 | 0 | 0 | 28 | 4 | 13 | 0 | 28 | 0.235294 | 1 | 23.52941 | 100 | 0.102879 | 0.033651 | 0.436938 | 0 | 1 | 1 |

Supplementary table S8:

Data table to show analysis of reports of oral swabs as sample type.

| Study | Participants | Comparison | TP | FN | FP | TN | Sens | Spec | Sens_Percentage | Spec_Percentage | SE_sens | LCI_sens | UCI_sens | SE_spec | LCI_spec | UCI_spec |
| --- | --- | --- | --- | --- | --- | --- | --- | --- | --- | --- | --- | --- | --- | --- | --- | --- |
| 2015_Wood_PCR | TB clinic patients | GeneXpert positive on sputum | 18 | 2 | 0 | 0 | 0.9 | NA | 90 | NA | 0.067082 | 0.768519 | 1 | NA | NA | NA |
| 2019_Luabeya_PCR | TB clinic patients | GeneXpert MTB/RIF plus MGIT | 49 | 10 | 6 | 65 | 0.830508 | 0.915493 | 83.05085 | 91.5493 | 0.048845 | 0.734772 | 0.926245 | 0.03301 | 0.850793 | 0.980192 |
| 2019_Mesman_Xpert TB/Rif | TB clinic patients | MGIT sputum culture | 15 | 18 | 0 | 0 | 0.454545 | NA | 45.45455 | NA | 0.086678 | 0.284656 | 0.624435 | NA | NA | NA |
| 2019_Nicol_PCR | Suspected pulmonary TB | IS Xpert MTB/Rif or sputum culture positive | 17 | 23 | 22 | 103 | 0.425 | 0.824 | 42.5 | 82.4 | 0.078162 | 0.271802 | 0.578198 | 0.034062 | 0.757239 | 0.890761 |
| 2020_Flores_PCR | Suspected pulmonary TB | Bacteriologically confirmed | 5 | 19 | 5 | 259 | 0.208333 | 0.981061 | 20.83333 | 98.10606 | 0.082898 | 0.045853 | 0.370814 | 0.008389 | 0.964617 | 0.997504 |
| 2020_Mesman_PCR | Culture confirmed TB | Culture positive | 63 | 60 | 0 | 0 | 0.512195 | NA | 51.21951 | NA | 0.04507 | 0.423858 | 0.600532 | NA | NA | NA |
| 2020_Molina-Moya_PCR | Clinical diagnosis presumptive TB | MTBC culture | 29 | 51 | 38 | 148 | 0.3625 | 0.795699 | 36.25 | 79.56989 | 0.053746 | 0.257157 | 0.467843 | 0.029563 | 0.737755 | 0.853643 |
| 2021_Ealand_MTBC g DNA | Young children less than equal 5 clinically diagnosed with TB | Gastric Aspirate culture/GeneXpert | 4 | 2 | 7 | 22 | 0.666667 | 0.758621 | 66.66667 | 75.86207 | 0.19245 | 0.289464 | 1 | 0.079463 | 0.602874 | 0.914368 |
| 2021_Ealand_Spogliotyping | | Gastric Aspirate culture/GeneXpert | 2 | 4 | 12 | 17 | 0.333333 | 0.586207 | 33.33333 | 58.62069 | 0.19245 | -0.04387 | 0.710536 | 0.091457 | 0.406951 | 0.765463 |
| 2021_Ealand_Auramine smear | | Gastric Aspirate culture/GeneXpert | 3 | 3 | 20 | 9 | 0.5 | 0.310345 | 50 | 31.03448 | 0.204124 | 0.099917 | 0.900083 | 0.085909 | 0.141963 | 0.478727 |
| 2021_Ealand_Culture | | Gastric Aspirate culture/GeneXpert | 0 | 6 | 0 | 29 | 0 | 1 | 0 | 100 | 0 | 0 | 0 | 0 | 1 | 1 |
| 2021_Song_TB-LAMP | > 16 years sypmtoms suggestuve pulmonary TB | MGIT | 33 | 5 | 8 | 55 | 0.868421 | 0.873016 | 86.84211 | 87.30159 | 0.054836 | 0.760942 | 0.9759 | 0.041948 | 0.790797 | 0.955235 |
| 2021_Wood_qPCR | Suspected TB | Culture | 43 | 4 | 12 | 44 | 0.914894 | 0.785714 | 91.48936 | 78.57143 | 0.040702 | 0.835117 | 0.99467 | 0.054832 | 0.678243 | 0.893185 |
| 2021_Wood_Culture | Suspected TB | Sputum culture | 82 | 50 | 0 | 9 | 0.621212 | 1 | 62.12121 | 100 | 0.042221 | 0.538458 | 0.703966 | 0 | 1 | 1 |
| 2022_LaCourse_qPCR | Suspected TB | Culture positive | 12 | 7 | 16 | 65 | 0.631579 | 0.802469 | 63.15789 | 80.24691 | 0.110665 | 0.414676 | 0.848482 | 0.044237 | 0.715764 | 0.889174 |
| 2022_Andama_Xpert Ultra | Adults cough for greater than 2 weeks | Xpert Ultra/Solid and MGIT culture | 42 | 16 | 0 | 125 | 0.724138 | 1 | 72.41379 | 100 | 0.058687 | 0.609111 | 0.839165 | 0 | 1 | 1 |
| 2022_Kang_SLIM assay | Clinically suspected of active PTB | Culture positive (sputum, BAL) | 64 | 35 | 40 | 133 | 0.646465 | 0.768786 | 64.64646 | 76.87861 | 0.048048 | 0.552291 | 0.740638 | 0.032054 | 0.70596 | 0.831613 |
| 2022_Shapiro_PCR | Adult over 16 | Sputum Xpert Ultra/Culture | 25 | 39 | 2 | 65 | 0.390625 | 0.970149 | 39.0625 | 97.01493 | 0.060986 | 0.271092 | 0.510158 | 0.02079 | 0.9294 | 1 |
| 2022_Cox_Xpert Ultra | <15 presumptive PTB | Induced sputum Xpert and culture | 20 | 70 | 1 | 200 | 0.222222 | 0.995025 | 22.22222 | 99.50249 | 0.043823 | 0.13633 | 0.308115 | 0.004963 | 0.985298 | 1 |

Supplementary table S9:

Other sample types

| Study | Participants | Method.of.testing | Comparison | TP | FN | FP | TN | Sens | Spec | Sens_Percentage | Spec_Percentage | SE_sens | LCI_sens | UCI_sens | SE_spec | LCI_spec | UCI_spec |
| --- | --- | --- | --- | --- | --- | --- | --- | --- | --- | --- | --- | --- | --- | --- | --- | --- | --- |
| Mouth wash |  |  |  |  |  |  |  |  |  |  |  |  |  |  |  |  |  |
| 1955_Rogers | Proven or suspected TB | Digestion, brought to 100ml and 10ml filtered | 24 hour sputum culture | 17 | 6 | 5 | 15 | 0.73913 | 0.75 | 73.91304 | 75 | 0.091561 | 0.559672 | 0.918589 | 0.096825 | 0.560224 | 0.939776 |
| 1955_Rogers | |  | Gastric aspirate | 1 | 2 | 0 | 88 | 0.333333 | 1 | 33.33333 | 100 | 0.272166 | -0.20011 | 0.866778 | 0 | 1 | 1 |
| 1994_Evans | Active TB and controls | Lysed RNA PCR | Broncoscopy/Sputum culture | 5 | 1 | 3 | 10 | 0.833333 | 0.769231 | 83.33333 | 76.92308 | 0.152145 | 0.535129 | 1 | 0.116855 | 0.540196 | 0.998266 |
| 2009_Davis | Outpatients and inpatients | PCR sec1 | Sputum culture | 55 | 20 | 6 | 46 | 0.733333 | 0.884615 | 73.33333 | 88.46154 | 0.051063 | 0.63325 | 0.833416 | 0.044305 | 0.797778 | 0.971453 |
| Saliva |  |  |  |  |  |  |  |  |  |  |  |  |  |  |  |  |  |
| 2015_Gonzalez | Adult suspected TB | Culture | Sputum culture | 28 | 4 | 0 | 0 | 0.875 | NA | 87.5 | NA | 0.058463 | 0.760412 | 0.989588 | NA | NA | NA |
| 2022_Byanyima | Xpert Ultra positive for TB on sputum | Xpert Ultra | Sputum culture | 70 | 8 | 2 | 1 | 0.897436 | 0.333333 | 89.74359 | 33.33333 | 0.034352 | 0.830106 | 0.964766 | 0.272166 | -0.20011 | 0.866778 |
| Other |  |  |  |  |  |  |  |  |  |  |  |  |  |  |  |  |  |
| 2003_Eguchi | Patients with TB | Ogawa egg medium | Sputum culture | 13 | 62 | 0 | 0 | 0.173333 | NA | 17.33333 | NA | 0.043709 | 0.087663 | 0.259004 | NA | NA | NA |
| 2003_Eguchi | Patients with TB | PCR | Sputum culture | 51 | 1 | 0 | 0 | 0.980769 | NA | 98.07692 | NA | 0.019045 | 0.943441 | 1 | NA | NA | NA |
| 2012_Palakuru | Pulmonary TB | Nested PCR | AFB positive sputum | 23 | 2 | 0 | 0 | 0.92 | NA | 92 | NA | 0.054259 | 0.813653 | 1 | NA | NA | NA |

SF1:

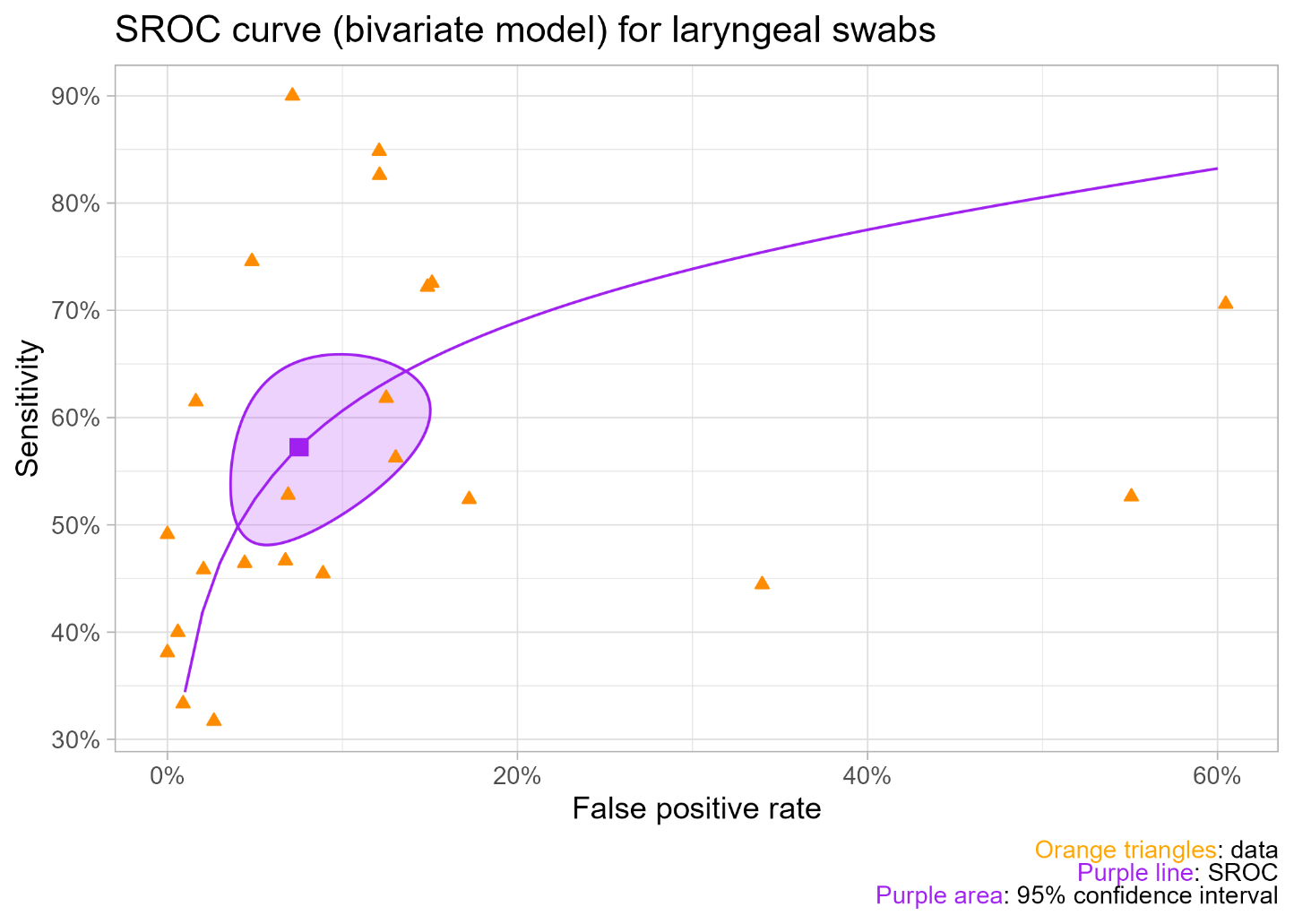

SF2:

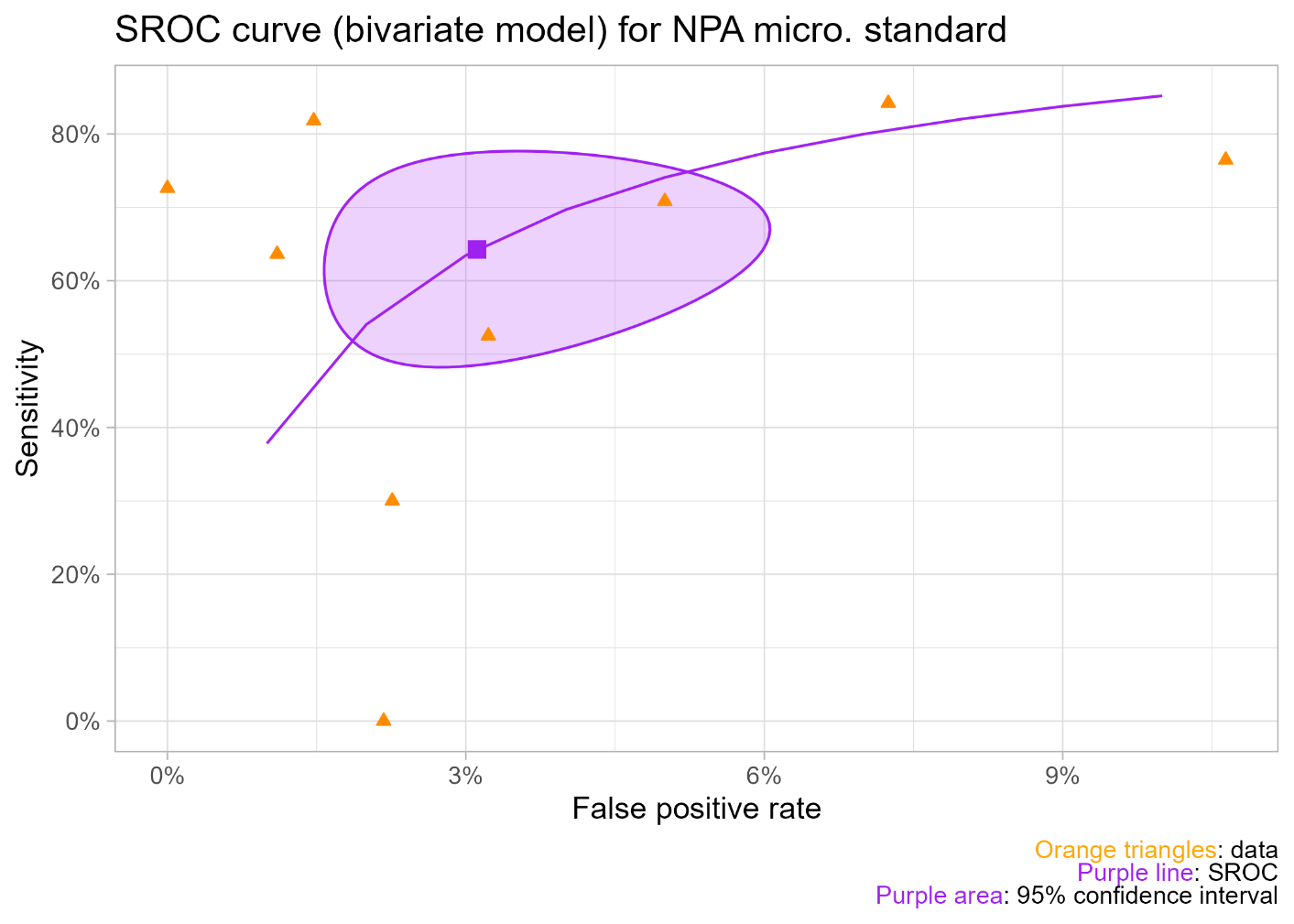

SF3:

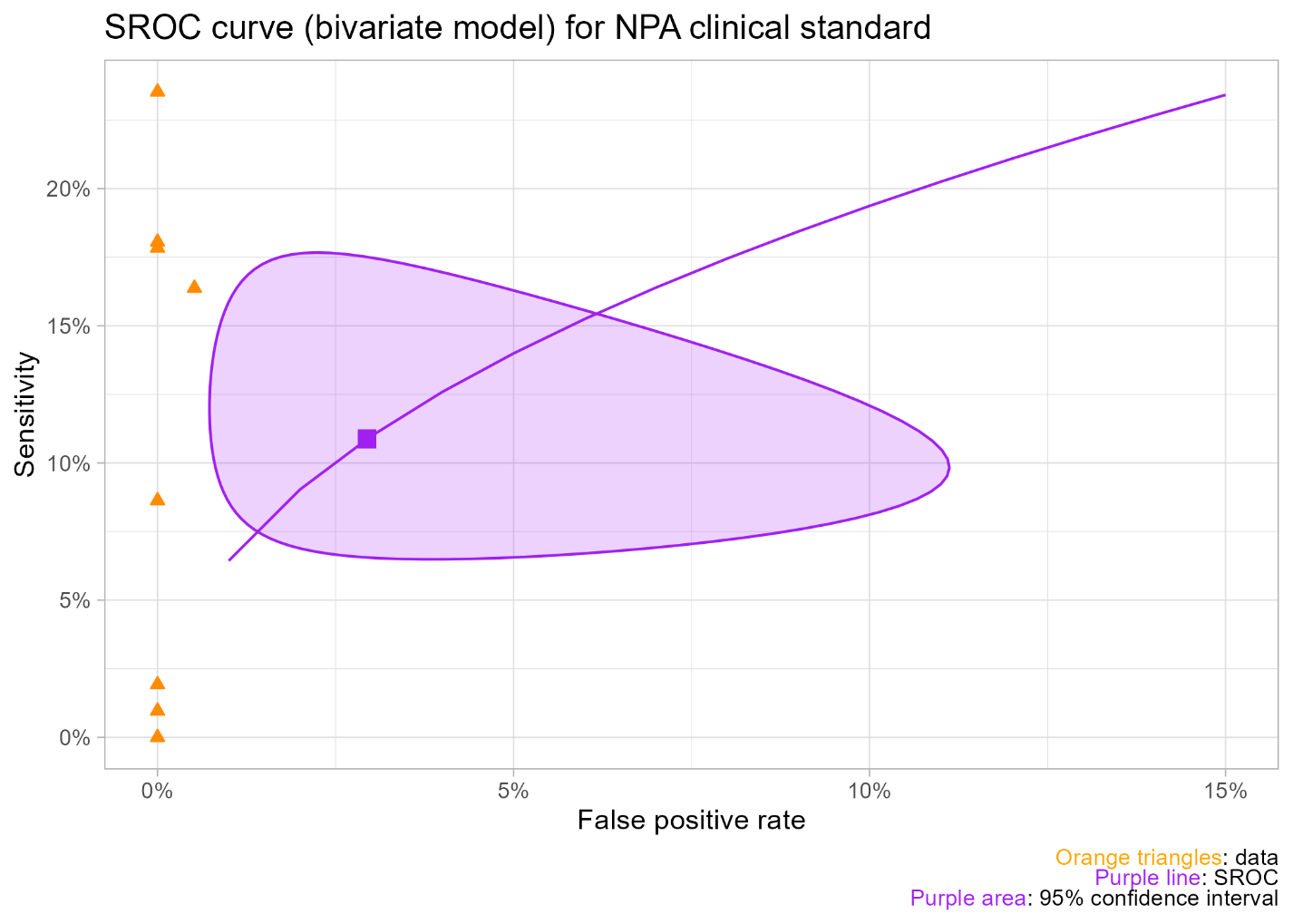

SF4:

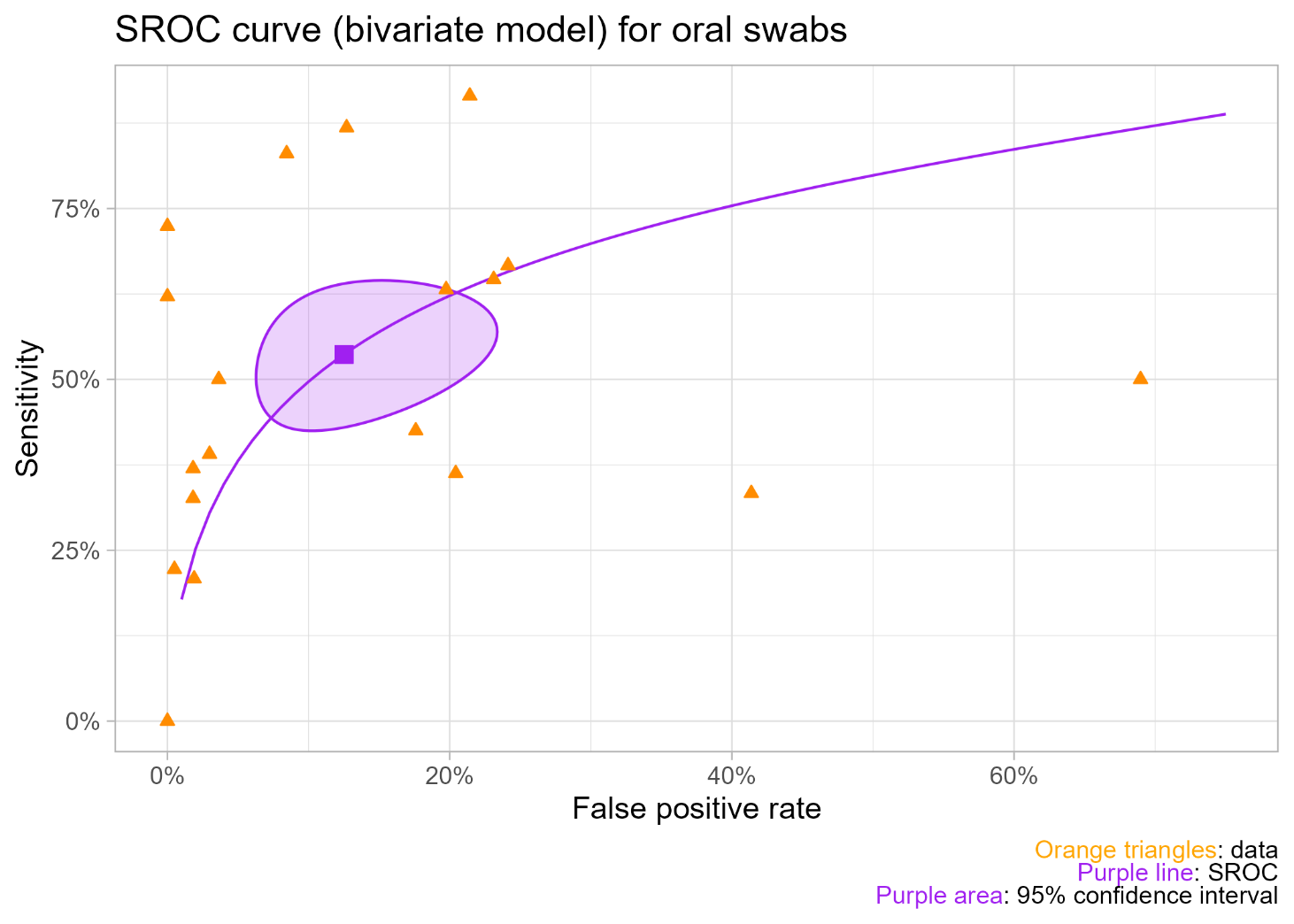

SF5: Bias of individual studies presented via each domain question and overall rating.

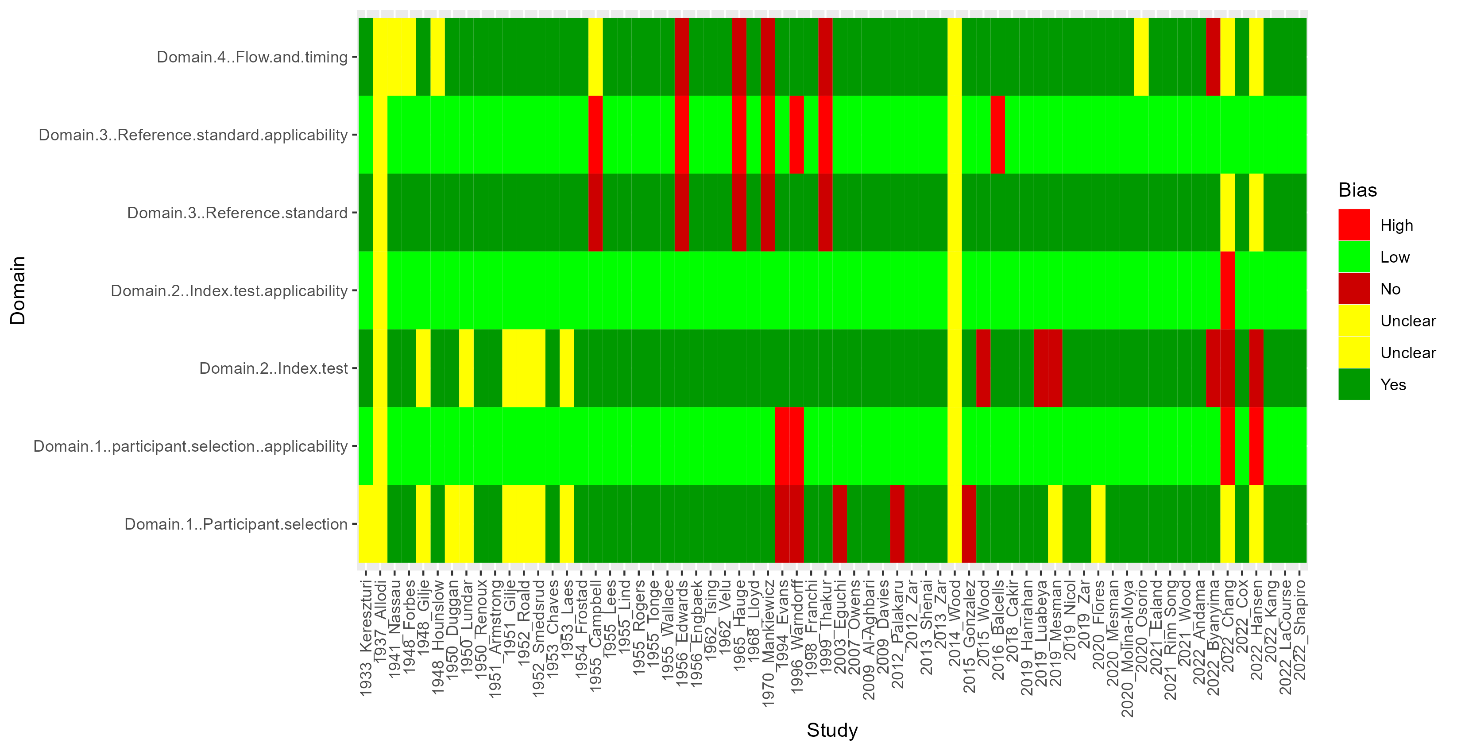

SF6: Bias of individual studies presented by domain.

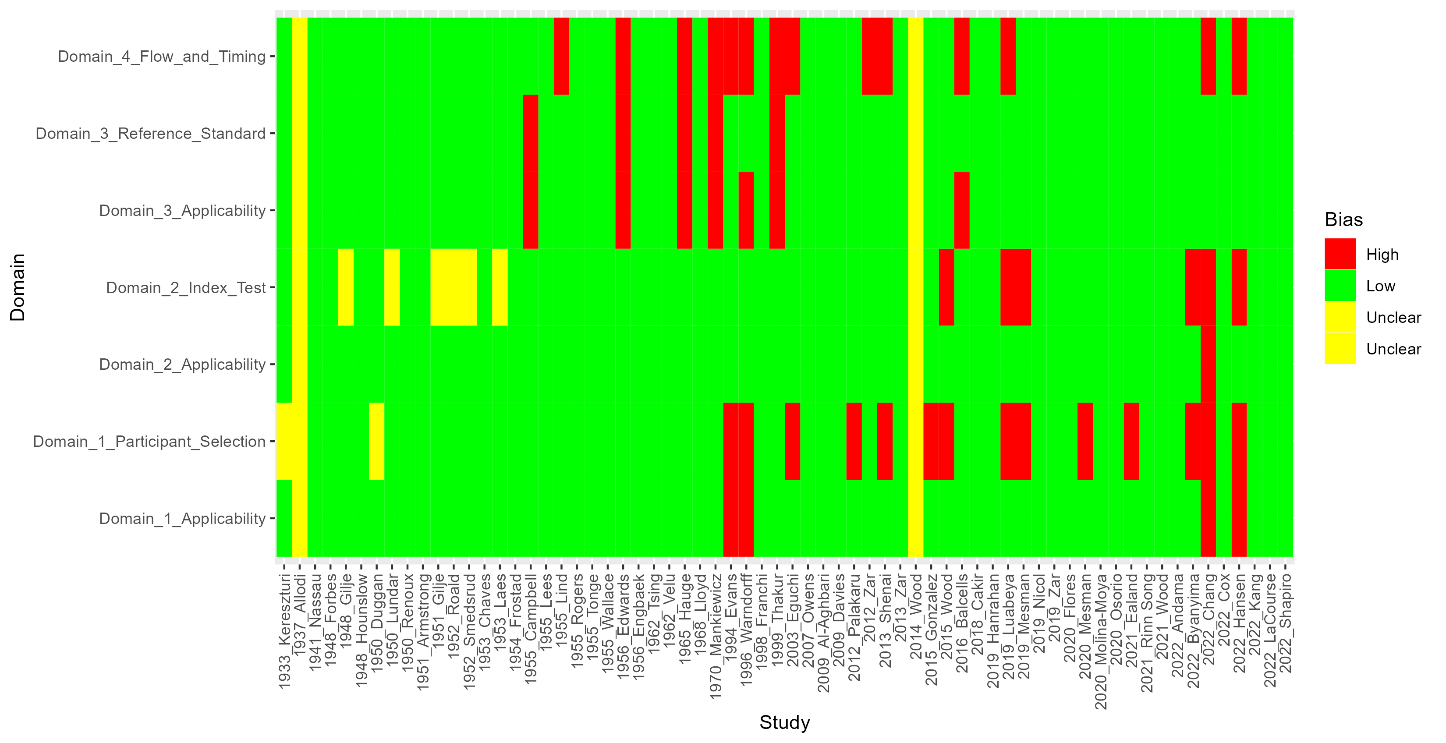
